# Tissue informative cell-free DNA methylation sites in amyotrophic lateral sclerosis

**DOI:** 10.1101/2024.04.08.24305503

**Authors:** C. Caggiano, M. Morselli, X. Qian, B. Celona, M. Thompson, S. Wani, A. Tosevska, K. Taraszka, G. Heuer, S. Ngo, F. Steyn, P. Nestor, L. Wallace, P. McCombe, S. Heggie, K. Thorpe, C. McElligott, G. English, A. Henders, R. Henderson, C. Lomen-Hoerth, N. Wray, A. McRae, M. Pellegrini, F. Garton, N. Zaitlen

## Abstract

Cell-free DNA (cfDNA) is increasingly recognized as a promising biomarker candidate for disease monitoring. However, its utility in neurodegenerative diseases, like amyotrophic lateral sclerosis (ALS), remains underexplored. Existing biomarker discovery approaches are tailored to a specific disease context or are too expensive to be clinically practical. Here, we address these challenges through a new approach combining advances in molecular and computational technologies. First, we develop statistical tools to select tissue-informative DNA methylation sites relevant to a disease process of interest. We then employ a capture protocol to select these sites and perform targeted methylation sequencing. Multi-modal information about the DNA methylation patterns are then utilized in machine learning algorithms trained to predict disease status and disease progression. We applied our method to two independent cohorts of ALS patients and controls (n=192). Overall, we found that the targeted sites accurately predicted ALS status and replicated between cohorts. Additionally, we identified epigenetic features associated with ALS phenotypes, including disease severity. These findings highlight the potential of cfDNA as a non-invasive biomarker for ALS.

## 1 Introduction

Cell-free DNA (cfDNA) is an emerging biomarker or biomarker candidate for multiple diseases, as it originates from dying tissues and can be non-invasively measured through a blood draw. CfDNA has been used in the detection of cancer,^1–3^ to identify fetal genetic abnormalities,^4,5^ to screen for infectious diseases,^6,7^ and to predict pregnancy complications.^8^ One underexplored domain for cfDNA, however, is in neurodegenerative disease. Biomarkers for neurodegenerative diseases are critically needed for improving patient care and evaluating the efficiency of clinical trials.^9^ While the application of cfDNA to neurodegeneration is nascent, our previous work,^10^ along with the work of others,^11–13^ has shown alterations in the cell-free DNA and RNA of patients with neurodegeneration relative to healthy controls.

Here, we build upon this work with a novel approach to epigenetic cfDNA biomarker development, applied to a large cohort of amyotrophic lateral sclerosis (ALS) patients. A limitation of whole genome epigenetic approaches is that the cost to achieve high sequencing coverage^14^ is currently too expensive to be routinely applicable in clinical settings.^2,15^ High sequencing coverage, however, is needed since certain cfDNA fragments may only be present in low quantities, which could be missed by shallow sequencing.^16^ Furthermore, many methylation sites are not variable,^17^ limiting their value in biomarker development.

To address these limitations, previous work has successfully used DNA methylation capture^18^ to enrich for only relevant genomic regions, which can reduce sequencing costs while maintaining high coverage. Examples of DNA methylation capture in cfDNA applications include an approach to classify cancer types and to predict whether a patient develops preeclampsia.^3,8,19^ A limitation of existing approaches, however, is that they are often optimized for a specific disease context. To the best of our knowledge, methylation capture of cfDNA has not yet been adapted for use in neurodegenerative disease.

In this work, we developed an algorithm to identify regions of the epigenome that are informative for the presence of a tissue in the cfDNA. These regions can be used to learn about tissue death in a range of diseases, including neurodegeneration. We then developed algorithms that leveraged differences in the methylation of these tissue informative sites to classify patients by disease status based on their epigenetic cfDNA profile. Our methodology can be used to characterize the contribution of diverse tissues to ALS cfDNA, leading to a multidimensional picture of disease.

We applied this technology to two independent cohorts from The University of Queensland in Brisbane, Australia (UQ) and the University of California at San Francisco, United States (UCSF) comprising a total of 192 cfDNA samples from ALS patients, healthy controls, and patients with other neurological diseases. Together, these cohorts represent the largest application of cfDNA in the study of ALS to date. Consistent with our previous research, we found significantly elevated cfDNA concentrations in ALS patients in both cohorts.^10^ Our machine learning model significantly predicted ALS disease status in both cohorts with high accuracy (UQ AUC=0.82, UCSF AUC=0.99). The model discriminated ALS cases from patients with other neurological diseases, such as frontotemporal degeneration. It also identified a previously unknown asymptomatic carrier of a pathogenic variant in *C9orf72,* which is the main genetic cause of ALS. Finally, we identified methylation sites associated with other ALS phenotypes, including disease severity. Together, these results suggest that epigenetic alterations in cfDNA are promising quantitative biomarker candidates for ALS, which can be used to non-invasively study the impacts of neurodegeneration.

## 2 Results

### 2.1 Overview of approach

The approach was composed of four steps. First, we analyzed published whole-genome bisulfite sequencing (WGBS) tissue data to identify methylation sites with distinct patterns in a tissue of interest. We call these sites tissue-informative markers (TIMs). Previously published cfDNA WGBS data from diverse disease contexts was used to screen candidate TIMs for those actually observed in cfDNA (See Methods). We then designed methylated and unmethylated probes complementary to these regions (Fig. 1a). Next, cfDNA from our two cohorts was extracted (Fig. 1b) and underwent methylation profiling on the TIM-enriched cfDNA (Fig. 1c). Lastly, we analyzed the methylation status of the targeted regions and developed statistical and machine learning approaches to learn about the disease status of the ALS patients and controls (Fig. 1d).

**Figure 1:**
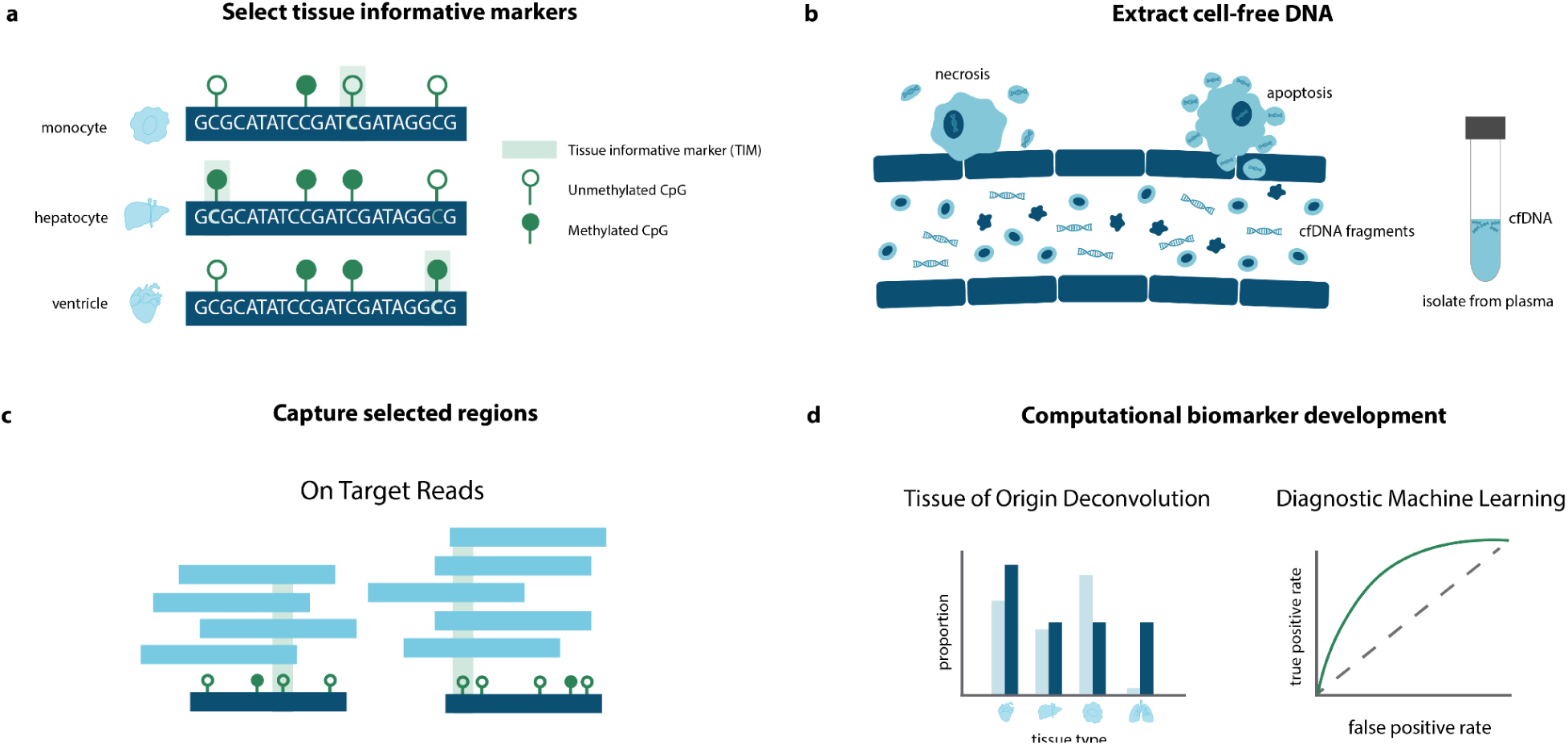
Overview of epigenetic cfDNA biomarker development approach. **(a)** Firstly, tissue informative markers (TIMs) were selected using WGBS data to capture CpG sites that were hypermethylated or hypomethylated in a tissue of interest. **(b)** Next, cfDNA was extracted from the blood plasma of ALS cases and controls. **(c)** The cfDNA was bisulfite-treated, hybridized to capture probes, designed as complementary to TIMs, and then sequenced. Some off-target reads were also captured. **(d)** Using computational approaches, we analyzed the tissue of origin of the cfDNA samples and performed machine learning to identify features of ALS.

### 2.2 Cohort characteristics

Our approach was applied to participants (n=192) who were recruited between 2018 and 2021 from two independent university-affiliated neurology clinics at UCSF and UQ (Table 1). The Revised El Escorial diagnostic criteria^20^ were used to classify cases (See Methods). Cases were composed of two groups of patients, those who had likely or probable ALS according to the criteria (referred to here as “ALS”), and those classified as possible ALS or primary lateral sclerosis (PLS) (referred to here as “PLS”), which is a related motor neuron disease.^21,22^

**Table 1:**
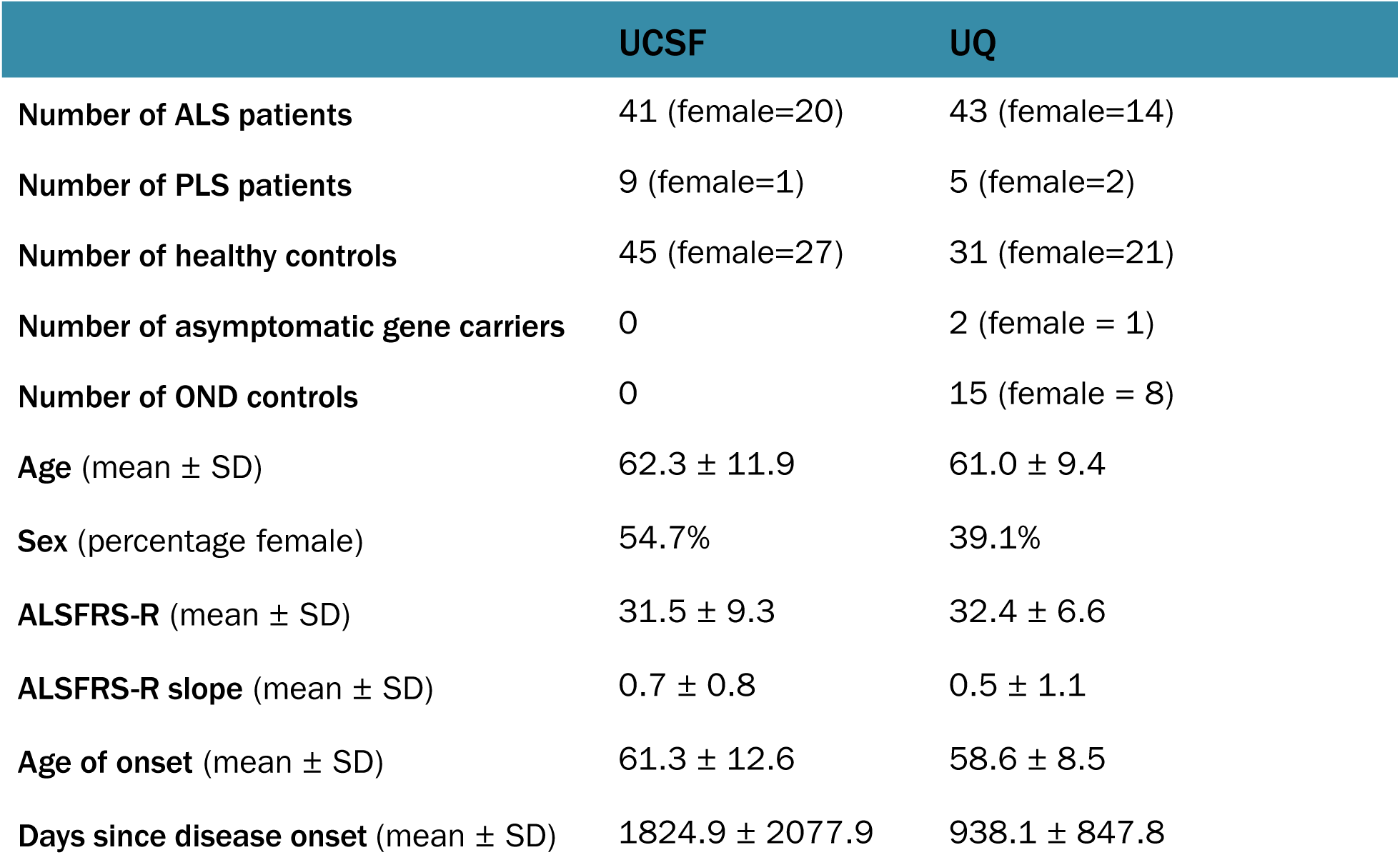
Clinical characteristics. The clinical and demographic characteristics per cohort. The number of total patients is shown, and the number of female patients is shown in parentheses.

The UCSF cohort comprised 42 ALS cases, 9 PLS cases, and 45 healthy age-matched controls consisting of unrelated partners or carers. At UQ, a total of 48 cases were enrolled (N=43 ALS and N=5 PLS). Forty-eight UQ controls were enrolled, consisting of both unrelated partners/carers (N=32) and patients with other neurological diseases (OND) (N=15). The UQ OND samples included a cross-section of neurological conditions, including diseases that share pathophysiology with ALS, like frontotemporal degeneration,^23^ and other neurodegenerative diseases like Alzheimer’s disease (Table 2). Therefore, the UQ cohort represented a challenging real-world scenario for ALS biomarker development.

**Table 2:** Other neurological disease patients. For each of the controls with other neurological diseases in the UQ cohort, the type of neurological disease (if known) and the number of patients with that disease.

| Disease Type | UQ OND Controls |
| --- | --- |
| Alzheimer's disease | 5 |
| Progressive supranuclear palsy | 3 |
| Frontotemporal degeneration | 2 |
| Other neurological disease | 1 |
| Parkinson's dementia (Lewy Body disease) | 1 |
| Corticobasal syndrome | 1 |
| Semantic dementia | 1 |
| Dementia with Lewy bodies | 1 |

There was heterogeneity of disease characteristics within and between cohorts. Both the UCSF and UQ cases had overlapping distributions in terms of age of onset, defined as the date the first ALS symptom was observed (Fig. 2a). For each cohort, ALS severity was measured using the ALS Functional Rating Scale-Revised (ALSFRS-R)^24^ at the time of cfDNA collection, which is a qualitative measure of physical functioning on a scale from 0 (not functioning) to 48 (high functioning). The change in ALSFRS-R between visits, referred to as ALSFRS-R slope, was also calculated as a metric of disease progression. We found that cohorts were similar in the distribution of ALSFRS-R and ALSFRS-R slope, although the UCSF had slightly more progressed cases (Fig 2b). UQ samples had higher forced vital capacity (FVC) (t-test p-value=4.5×10^-5^), which is a measure of lung function, where a higher value indicates better function (Fig. 2c). The two cohorts were also similar in the distribution of days between cfDNA collection and symptom onset (Fig. 2d). We noted that patients in the UCSF cohort were slightly older (UQ mean age: 61.45 ± 8.17, UCSF mean age: 66.33 ± 9.96)) and that the UCSF cohort also contained patients from a larger variety of self-reported racial and ethnic (SIRE) backgrounds (Fig. S1b-d).

**Figure 2:**
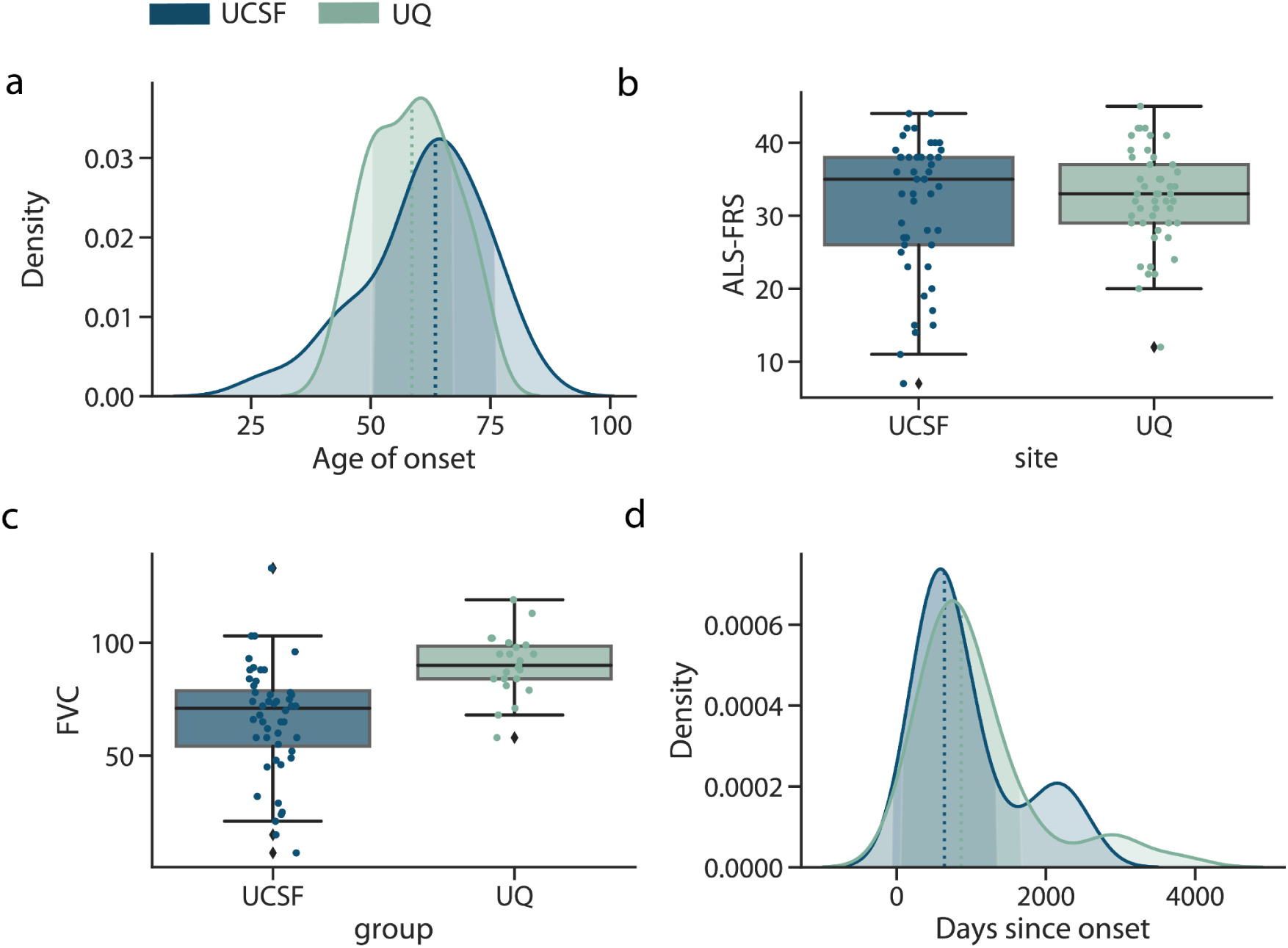
Cohort demographic and clinical characteristics. For the UQ (n=43) and UCSF (n=42) ALS patients, **(a)** the distribution of the age of onset of ALS disease symptoms, where the dotted line indicates the median age of onset, **(b)** patient ALSFRS-R scores **(c )** FVC, and **(d)** the number of days between cfDNA collection and date ALS symptoms were observed. In the box plots, the center line of the box indicates the mean, the outer edges of the box indicate the upper and lower quartiles, and the whiskers indicate the maxima and minima of the distribution. Each dot indicates an individual.

### 2.3 Selecting tissue informative markers

After collecting cfDNA, we turned to selecting methylation sites that are variable between tissues. In our previous work,^10^ we introduced the concept of tissue informative markers (TIMs) as a method to identify methylation sites that vary between tissues and cell types. Briefly, a TIM is a site that is either hyper- or hypo-methylated relative to the average methylation proportion of all other tissues at that site (Fig. 3a) (See Methods).

**Figure 3:**
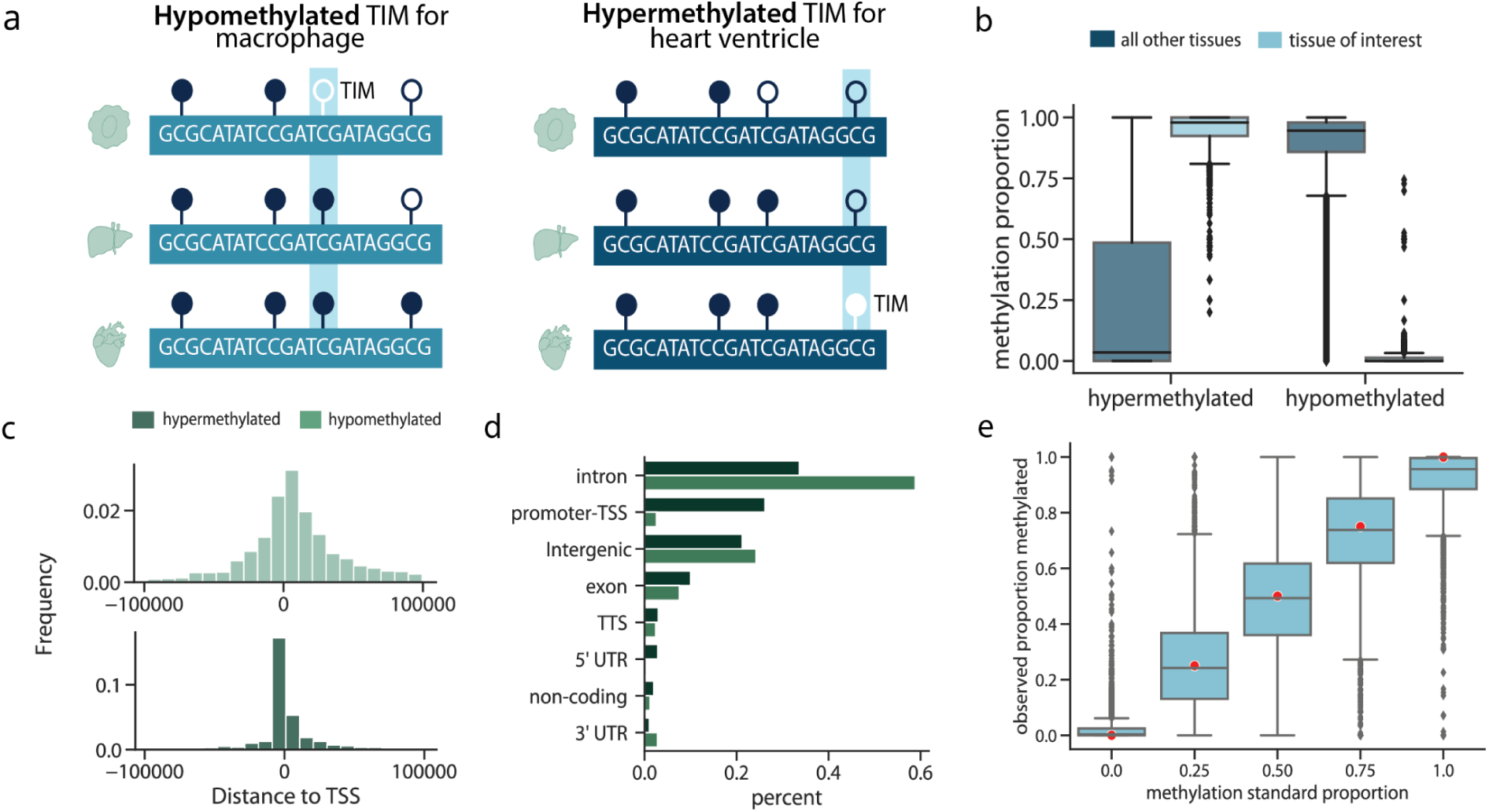
Capture panel design. **(a)** The panel was designed to capture both hypomethylated TIMs, which were CpG sites that were less methylated in a tissue of interest relative to other tissues, and hypermethylated TIMs, which were designed to capture sites more methylated in a tissue of interest than other tissues. **(b)** The methylation proportion of reference tissues at either the site the TIM was selected for, or all other tissues. **(c)** The distance hyper- or hypo-methylated TIMs are from the transcription start site of a gene. **(d)** The number of hyper- and hypo-methylated TIMs in different genomic regions. **(e)** For samples where the true genome-wide methylation proportion was between 0.0 and 1.0 (red dots), the observed methylation proportion after capture and sequencing. For all box plots, the center line of the box indicates the mean, the outer edges of the box indicate the upper and lower quartiles, and the whiskers indicate the maxima and minima of the distribution. Each dot indicates an individual.

To find TIMs, we used reference WGBS methylomes that were obtained from two public reference consortiums, ENCODE^25^ and Blueprint.^26^ For this work, we focused on CpG sites as candidate TIMs, as most non-CpG sites are not methylated in adult tissues.^27^ We selected approximately 300 TIMs for 19 tissues (Table 3), which were prioritized based on deconvolution results from our previous work^10^ and other recent works.^28,29^ These tissues included several hematopoietic cell types, organs, epithelium, and brain (Table 3). We applied several filtering criteria to enrich for CpG sites that appear in previously published WGBS cfDNA data.

**Table 3:** TIM selection design. Per tissue selected for capture, the number of hypermethylated TIMs selected, the number of hypomethylated TIMs selected, and the total number of final TIMs selected for capture.

| Tissue | Hypermethylated | Hypomethylated | Total |
| --- | --- | --- | --- |
| Adipose | 208 | 42 | 250 |
| Brain | 200 | 50 | 250 |
| Dendritic cell | 179 | 71 | 250 |
| Endothelial cell | 200 | 50 | 250 |
| Eosinophil | 83 | 167 | 250 |
| Erythroblast | 134 | 116 | 250 |
| Fibroblast | 200 | 50 | 250 |
| Heart left ventricle | 204 | 46 | 250 |
| Hepatocyte | 200 | 50 | 250 |
| Lung left lobe | 200 | 45 | 245 |
| Macrophage | 86 | 164 | 250 |
| Mammary | 200 | 50 | 250 |
| Megakaryocyte | 184 | 66 | 250 |
| Monocyte | 72 | 164 | 236 |
| Neutrophil | 66 | 184 | 250 |
| Placenta | 200 | 50 | 250 |
| Skeletal muscle | 200 | 50 | 250 |
| Small intestine | 200 | 50 | 250 |
| T-cell | 290 | 223 | 513 |

An important property of cfDNA is that their fragmentation patterns are non-random.^30–32^ cfDNA observed in blood generally are fragments approximately 160 base pairs long,^33^ suggesting that cfDNA fragments are protected from degradation in the blood by the presence of tightly associated histone proteins. Since DNA from compacted chromatin is more likely to be protected and methylated, we chose to select a greater number of TIMs per tissue that were hypermethylated (Table 3) (Fig. 3b).

After quality control (Methods), the final number of TIMS was 4,994. TIM sites were distributed throughout the genome (Fig. S2a). Hypermethylated TIMs were closer, on average, to transcription start sites and CpG Islands than hypomethylated TIMs (Fig. 3c; Fig. S2b). Since at a hypermethylated TIM, all other tissues are predominantly unmethylated, this observation is consistent with the role of unmethylated CpGs in facilitating transcription.^34^ Likewise, hypomethylated TIMs were more likely to be in intergenic and intronic regions (Fig. 3d), suggesting that in most tissues, these sites did not have a strong regulatory function. Together, this suggests that hypermethylated and hypomethylated TIMs offer complementary types of genomic information.

### 2.4 Capture panel sequencing and validation

After designing the probes, we performed several validation experiments to ensure that probes could accurately profile the methylation state of the chosen TIMs. First, we used universal methylated DNA standards to create mixtures where the CpG sites were methylated 0, 25, 50, and 100% of the time. We captured the synthetic DNA mixtures with the probes and performed high-throughput sequencing. For each DNA mixture, we estimated the proportion of the time the captured CpG was methylated. We found that the observed methylation was highly concordant with the true methylation proportion (Fig. 3e), suggesting that the probes were quantifying the methylation accurately.

Next, to examine how the capture panel might perform in real-world cfDNA scenarios, we validated the capture panel using sheared genomic DNA from blood (n=2), along with healthy cfDNA samples (n=3). After performing cell-type deconvolution with CelFiE,^10^ we found that the sheared blood samples were estimated to be primarily composed of white blood cells (Fig. S3a). The majority of cfDNA from healthy controls was also estimated to be originating from neutrophils and lymphocytes, consistent with published research (Fig. S3b).^35^

Lastly, we extracted cfDNA from a healthy control before and after vigorous exercise to examine the ability of the panel to measure tissue-specific changes in biological state. After capture and sequencing, we performed deconvolution of these two cfDNA samples. We found that cfDNA originating from neutrophils increased in the sample taken after exercise (Fig. S3c), consistent with a recent report^36^ studying the effect of exercise on cfDNA composition. Together, these experiments demonstrate that our approach for targeting TIMs can correctly capture the methylation state of cfDNA and measure relevant tissue of origin effects.

### 2.5 cfDNA capture from ALS cases and controls

We next turned to examining the cfDNA epigenome of our disease cohorts. cfDNA was extracted from the blood plasma of cases and controls from both UQ and UCSF patients. We first confirmed our previous finding^10^ of an increased concentration of cfDNA in the plasma of ALS patients relative to controls after correcting for age, sex, and SIRE (Fig. 4a) (logistic regression UQ: log odds ratio=7.5×10^-3^, p-value=1.8×10^-2^, UCSF: log odds ratio=2.4×10^-2^, p-value=6.0×10^-3^). Interestingly, cfDNA was also elevated in ALS patients relative to the OND controls (logistic regression log odds ratio=1.6×10^-2^, p-value=3.6×10^-2^), which had overall low levels of cfDNA. This suggests that the cfDNA generative processes of apoptosis and necrosis might differ between ALS and other types of neurological diseases.

**Figure 4:**
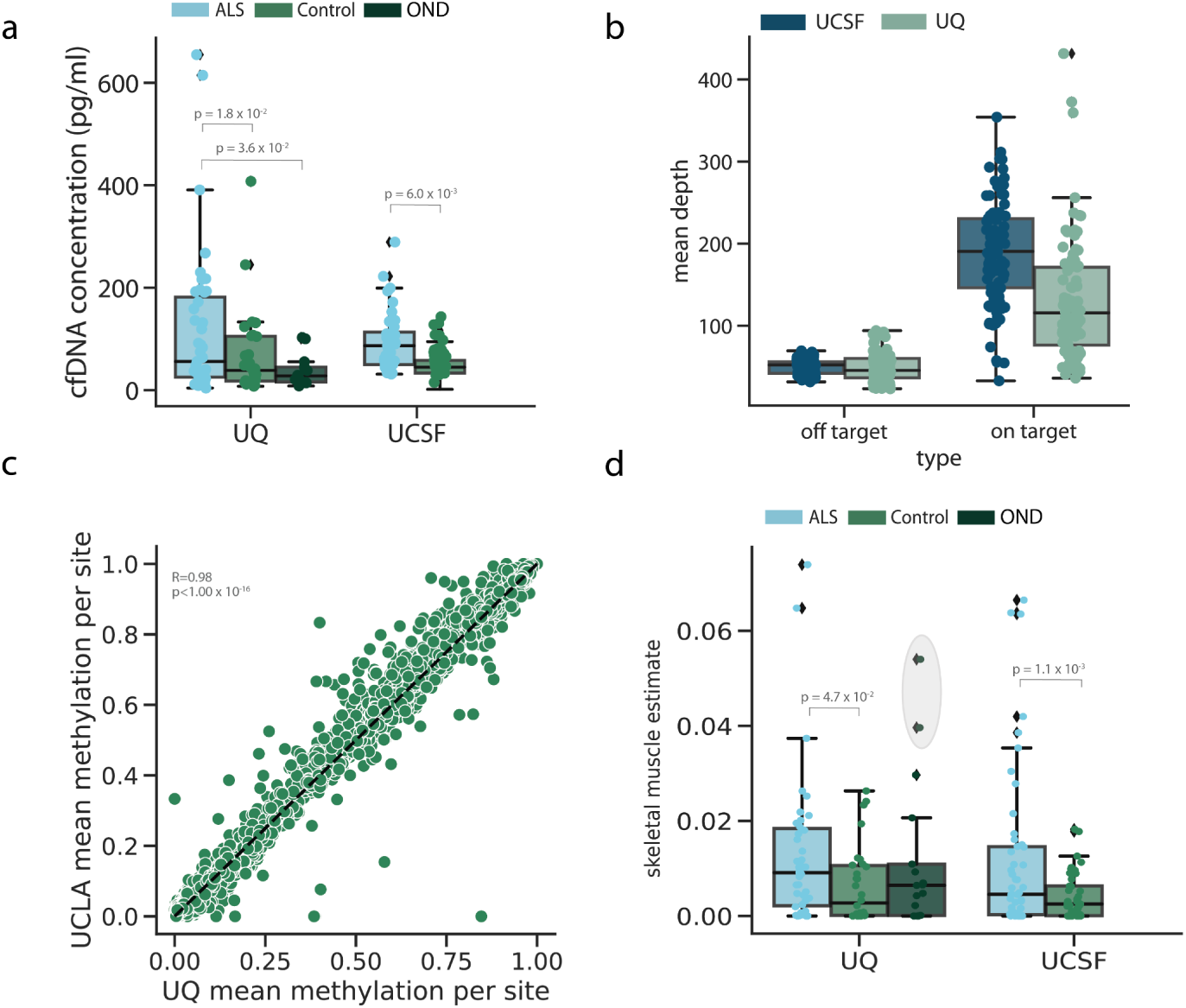
Capture panel performance on cfDNA data. **(a)** The starting cfDNA concentration of ALS patients and controls for each cohort, where each point represents one individual. **(b)** Coverage of the on-target and off-target CpG sites of each cohort, where each dot represents one sample. **(c)** Correlation between the UQ and UCSF methylation proportions at on-target sites. A single point represents a TIM. (**d**) The proportion of cfDNA from the controls and cases in each cohort that was estimated to originate from skeletal muscle. The grey shaded circle indicates outlier control individuals discussed in Section 2.6. For all box plots, the center line of the box indicates the mean, the outer edges of the box indicate the upper and lower quartiles, and the whiskers indicate the maxima and minima of the distribution. Each dot indicates an individual.

After quantifying the amount of cfDNA, we performed high-throughput methylation sequencing on the captured regions. Since bisulfite treatment can degrade the already low quantity of input DNA, cfDNA sequencing experiments are prone to high duplication.^16^ To address this, we used unique molecular identifiers (UMIs) to deduplicate reads. In total, after sequencing and deduplication, the average on-target coverage of UQ samples was 134 ± 166 reads per CpG and the average on-target coverage of UCSF samples was 195 ± 229 reads per CpG. The average methylation proportion at TIM sites was highly correlated between the two cohorts (Pearson’s R=0.98, p<1.0×10^-16^) (Fig. 4c).

We noted that UCSF samples had a higher percentage of on-target reads (Fig. S4), which likely contributed to differences in overall CpG read coverage. We also found that cfDNA starting concentration was a significant predictor of on target saturation after adjusting for total on target coverage (linear regression effect size=-1.7 × 10^-3^, p-value=9.0 × 10^-3^) (Fig. S4d-e).

### 2.6 Cell-type decomposition

Since TIMs were designed to be specific to a given tissue type, they can be used to estimate what tissues are contributing to the cfDNA in the context of neurodegeneration. To do this, we performed cfDNA cell-type decomposition with CelFiE.^10^ CelFiE is a supervised decomposition algorithm that is designed to work with methylation read count data and missing or noisy reference data. As input, CelFiE takes the TIM read count data for each cfDNA sample and estimates the proportion of the cfDNA mixture originating from the tissues in the reference dataset, along with a specified number of unknown tissues.

We ran CelFiE with two unknown components using the methylation proportion of the captured sites as input (Fig. S5a-b). As with our prior ALS study^10^, we observed elevated skeletal muscle in ALS patients in both cohorts relative to the healthy control samples (t-test p-value UCSF: 1.1×10^-3^, UQ: 4.7×10^-2^) (Fig. 5d). This is consistent with muscle atrophy that occurs as part of their disease. We then tested the remaining tissue estimates for association with ALS disease status. At nominal significance (p<0.05) CelFiE estimated a depletion of cfDNA originating from eosinophils in ALS cases (t-test p-value UCSF: 1.0×10^-2^, UQ: 9.3×10^-3^)(Fig. S5c-d). While a preliminary result,previous studies have observed changes in granulocyte counts in the whole blood of ALS patients^37,38^ and overall immune dysregulation is thought to be an important contributor to ALS etiology.^39^

**Figure 5:**
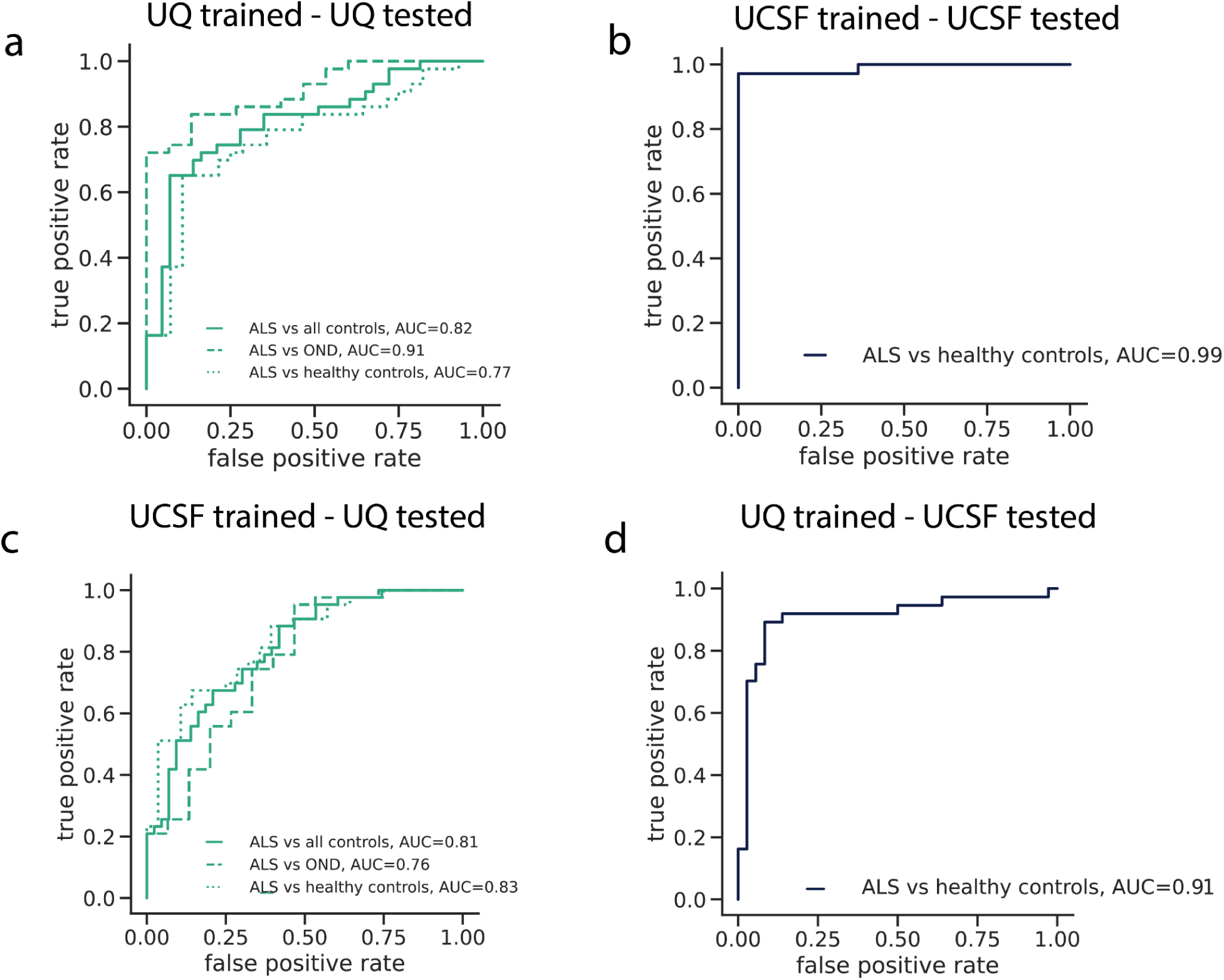
ALS disease classification with cfDNA epigenetic features. The false positive rate versus true positive rate for models trained and tested using CpG coverage, CpG methylation, and covariates as input features for **(a)** ten-fold cross-validation within UQ samples **(b)** ten-fold cross-validation within UCSF samples **(c)** trained on UCSF data and tested on UQ data, and **(d)** trained on UQ data and tested on UCSF data.

Interestingly, we observed two UQ control samples with unusually high skeletal muscle components (an estimated 5.4% and 3.9% of their total cfDNA sample) (Fig. 5d). One sample was an OND control with frontotemporal dementia, a disease that has substantial genetic and clinical overlap with ALS.^40^ The other sample was originally classified as a healthy control. However, after further investigation into their clinical records, this individual had both a parent and sibling with ALS. Genetic testing revealed that this individual also tested positive for a C9orf72 repeat expansion, which is the most common genetic cause of ALS,^41^ suggesting that the individual may be presymptomatic. Since the disease status of this patient was ambiguous, we reclassified them as OND.

### 2.7 Classification of ALS disease status

While muscle degeneration is a hallmark of ALS, it is not specific enough to serve as a diagnostic tool. Therefore, to further characterize the relationship between alterations in the cfDNA epigenome and disease, we developed a tissue-agnostic algorithm that utilized information from all TIM epigenetic profiles to predict whether a cfDNA sample was from an ALS patient or control. For these models, we did not consider PLS samples, but return to these samples in Section 2.9. Further models integrated all CpG sites, both on and off target (See Methods).

We trained an elastic net prediction model^42^ in four contexts to explore the generalizability of the results across the independent cohorts. First, a model was trained using ten-fold cross-validation within each cohort. Then, the transferability of the models was assessed by training a model on one cohort and applying it to the other. Since only the UQ cohort had OND and healthy controls, we combined the controls for this analysis, although we later examined the ability of the model to discriminate between the different sample types. Model parameters, including the elastic net mixing parameter, were selected by using a cross-model selection and averaging procedure within the training set.^42^ Non-penalized covariates included age at the time of cfDNA sampling, sex, SIRE, cfDNA concentration, and total cfDNA input. We evaluated model performance with area under the receiver operating characteristic curve (AUC) and by testing whether the predictions could significantly predict true case-control status using a logistic regression model that included covariates.

To best characterize the different types of information that TIMs can provide we explored two classes of features for the prediction model, the methylation proportion and the coverage of the TIMs. Coverage was included because cfDNA fragmentation is non-random (see Section 2.3); we therefore reasoned that CpG coverage may also be informative of disease status. In total, we trained models using CpG coverage only, CpG methylation proportion only, and a combination of both as input features.

Overall, we found that tissue informative epigenetic features could significantly predict ALS case-control status in both cohorts (Fig. 5, Fig. S6-7, Table 4). The best-performing model incorporated both TIM coverage and methylation features (Fig. 5). Within cohorts, the ten-fold cross-validated AUC was 0.82 within the UQ cohort (logistic regression odds ratio=2.34, p=2.32×10^-7^) and the UCSF AUC was 0.99 (logistic regression odds ratio=2.51, p-value<2.0×10^-16^). The models were more predictive than models trained using only covariate information (Fig. S8). Importantly, even though the model was not trained to distinguish between ALS cases and OND, the AUC was high for both UQ models (within UQ: AUC=0.91, UCSF-UQ: AUC=0.76).

**Table 4:**
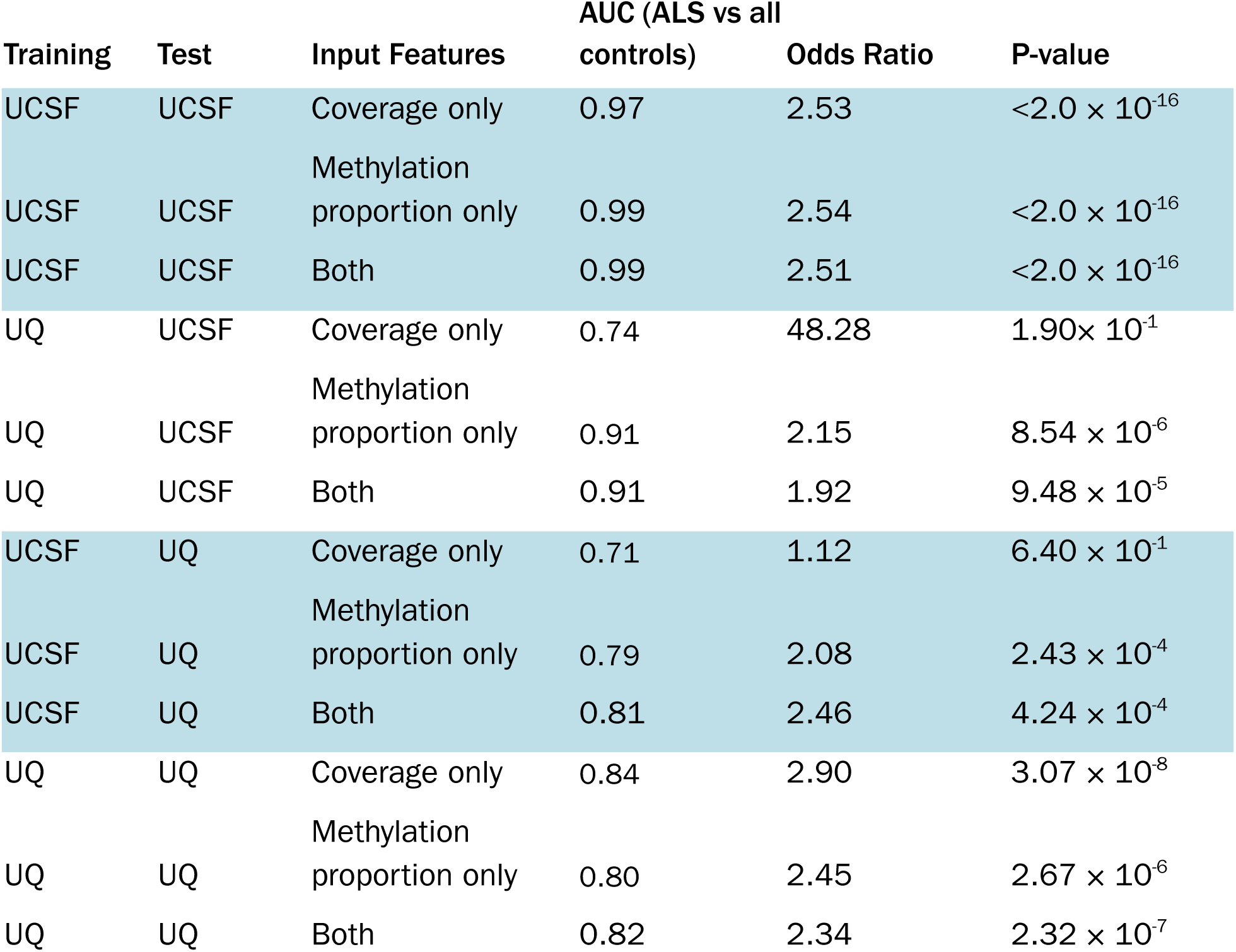
Binary prediction model performance. The AUC of predicting ALS vs all control samples for four models trained either within a cohort or trained in one cohort and tested on the remaining cohort. Models were trained with either only CpG coverage as input features, only CpG methylation, or both.

Models trained within one cohort replicated between cohorts. We noted that the prediction performance was higher for the UQ-trained and UCSF-tested model (AUC=0.91, logistic regression odds ratio=1.92, p=9.48 × 10^-5^) than the UCSF-trained model applied to the UQ samples (AUC=0.81, logistic regression odds ratio=2.46, p=4.24×10^-4^). Differences in model performance between cohorts were likely driven by a combination of factors, including cohort heterogeneity and technical variation. One likely contributing factor was the lower on-target coverage in the UQ cohort (Fig. 4d, Fig. S5). To test this, we randomly downsampled the number of reads in each UCSF cfDNA sample, which reduced effective on-target CpG coverage. Then, we re-ran the elastic net model within the UCSF cohort. We found that lower read coverage led to worse classification performance (Fig. S9), suggesting that on-target CpG coverage is an important factor in prediction accuracy.

Importantly, the predictive performance of the elastic net models was stronger than using the CelFiE skeletal muscle estimate alone (Fig. 4d). Indeed, models trained without any skeletal muscle TIMs, did not have reduced performance relative to the full model (Fig. S10), emphasizing the importance of combining information across tissue contributors.

We also noted that despite methylation proportion being the more common feature considered in epigenetic cfDNA studies, the models trained only using CpG coverage also significantly predicted case-control status (Table 4, Fig. S6). In fact, there was very similar performance within the UCSF cross-validated model (AUC=0.97) and the UQ cross-validated model (AUC=0.84). This suggests that disease-relevant information is contained in simply the observation of a given CpG in cfDNA sequencing data, providing an additional layer of information over the CpG methylation state alone. This information may be lost in other low-cost epigenetic assays, like methylation arrays, that only return methylation proportion values.

Lastly, we considered the elastic net models that incorporated off-target CpGs (UCSF total number of CpGs=32314, UQ total number of CpGs=49238). We found that the off-target models performed well (UCSF AUC=0.86, UQ AUC=0.76), even though this was a challenging setting as there were many more features than samples (Fig. S11). In future work, sites selected in these models could be chosen to refine TIM selection for capture panel development.

### 2.8 Biological significance of prediction features

Next, we sought to understand how different tissue informative sites contributed to predicting disease. An advantage of using a regularized regression model like an elastic net is that the model performs feature selection and assigns a higher weight, or absolute β value, to features that contribute more to accurately predicting the outcome. Features that do not contribute to the prediction will have an absolute β value near zero. Thus, to examine the overall contribution of different types of TIMs in making model predictions, we obtained the absolute β value for each TIM from an elastic net model trained on the entire UQ and UCSF cohorts (Fig. 5c and Fig. 5d). Then we examined how these values related to different characteristics of the TIMs.

We first analyzed whether TIMs selected for a given tissue type were more important in making predictions. As expected, skeletal muscle TIMs were highly important in making model predictions, especially for TIMs that were hypermethylated in skeletal muscle (Fig. 6a). Despite the importance of skeletal muscle TIMs, we noted that TIMs for every tissue type contributed to the model predictions (Fig. 6). This again highlights the contribution of multiple tissues in neurodegeneration and the possibility of designing disease-specific biomarkers. For example, T-cell TIMs were highly important (Fig. 6a), indicating that cfDNA originating from immune cell types may be relevant in ALS disease.

**Figure 6:**
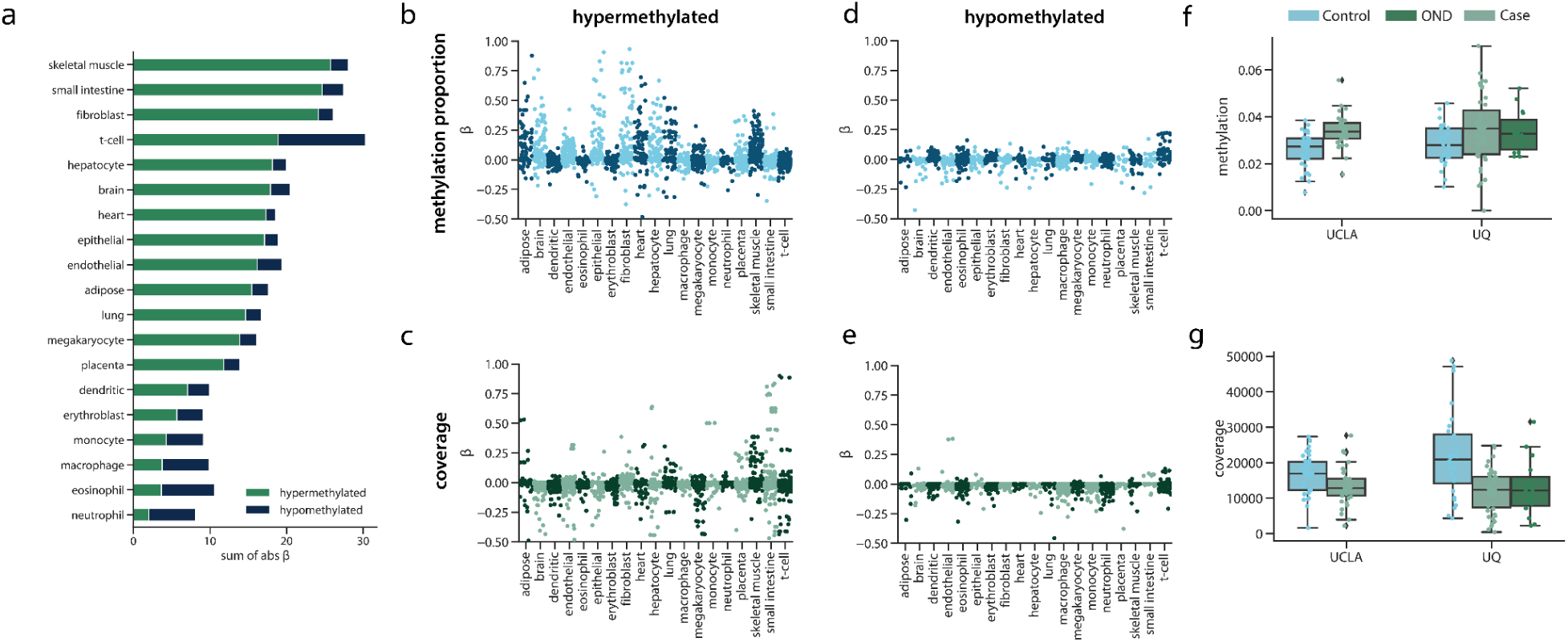
Features selected by the elastic net algorithm. **(a)** For each tissue the TIMs were selected for, and for the type of TIM, the total absolute β value. A larger absolute β sum indicated that the feature type contributed more to model predictions. The β values for the **(b)** methylation proportion and **(c)** the read coverage of individual TIMs selected to be hypermethylated, and the β values for the **(d)** methylation proportion and **(e)** read coverage of individual TIMs selected to be hypomethylated. **(f)** The methylation proportion of cases and controls for each cohort for a hypermethylated TIM in the *SHISA5* gene. **(g)** The read coverage of cases and controls for each cohort for a hypermethylated TIM located in the *XRCC6* gene. For all box plots, the center line of the box indicates the mean, the outer edges of the box indicate the upper and lower quartiles, and the whiskers indicate the maxima and minima of the distribution. Each individual dot indicates a cfDNA sample.

Overall, there were differences in the importance of each class of TIM. Hypermethylated TIMs generally had higher absolute β values than hypomethylated TIMs (Fig. 6b-e), which could be related to our previous observation that hypermethylated TIMs were more likely to be in promoter or genic regions (Fig. 3). We also observed that there were differences in the distribution of absolute β values of methylation proportion and coverage features. For example, while methylation proportion features for fibroblast and epithelial cells had high absolute β values, coverage features for these tissues had relatively low absolute β values (Fig. 6b-c). Instead, the coverage of TIMs for small intestine and T-cells were high, but close to zero as methylation proportion features. Together, this could mean that including both methylation proportion and coverage of tissue informative sites is useful for learning about disease in the context of cfDNA.

We next examined individual TIMs as an avenue for examining and generating hypotheses about individual epigenetic biomarker candidates. TIMs with a non-zero absolute β value were chosen for association with ALS case-control status, along with covariates and correcting for cohort. Multiple test correction was employed using false discovery rate at 10%. One of the most important methylation proportion features was a hypermethylated TIM selected for epithelium. We observed significantly increased methylation in ALS cases for this TIM (logistic regression odds ratio=14.09, q-value=8.06×10^-2^) suggesting that there was increased contribution from this gene in the cfDNA of ALS patients (Fig. 6e). This TIM was located in the promoter region of the *SHISA5* gene, which, along with *p53*, is involved in apoptosis.^43^ Additionally, *SHISA5* was found to be over-expressed in the spinal cord of ALS patients.^44^

We identified a similarly interesting hypermethylated TIM in the coverage features. While the TIM was selected for hepatocytes, it is in the *XRCC6* gene, which was highly expressed in many tissues in bulk RNA-seq from the Genotype-Tissue Expression (GTEx) Project.^45^ The TIM had significantly reduced coverage in ALS patients relative to controls (logistic regression odds ratio=-42.25, q-value=1.35 × 10^-2^) (Fig. 6f), and while it is difficult to infer directly from cfDNA alone, this result could suggest potential dysregulation of this gene in cases. *XRCC6* is involved in non-homologous end joining and DNA repair.^46^ Disruption of non-homologous end joining has been previously linked to aging and ALS.^47,48^

### 2.9 ALS disease phenotypes

To further explore the value of tissue-specific methylation sites as a potential biomarker, we developed models to predict ALS disease phenotypes. To do this, we trained three linear elastic net models to predict ALSFRS-R (n=78), ALSFRS-R slope (n=60), and FVC (n=57) with ten-fold cross-validation. We hypothesized that high-weight features from the case-control analysis would also be associated with ALS phenotypes, and so, we chose the top 1000 coverage and top 1000 methylation features with the highest absolute β as input for the models. Since case-only numbers were relatively low in each cohort, we meta-analyzed the two cohorts, adding a non-penalized covariate for each cohort in the analysis, along with age, sex, cfDNA concentration, total cfDNA input quantity, and SIRE. To specifically evaluate the performance of cfDNA features over covariates, we separately trained an additional three models using only covariates.

We found the models based on cfDNA epigenetic features significantly predicted ALSFRS-R (Fig.7a) (Pearson’s R=0.66, p-value=3.71×10^-9^). This was statistically significantly higher (p-value=1.85×10^-5^) than the predictions from the model trained only on covariates (Pearson’s R=0.49, p-value=5.81×10^-5^). We found that the high predictive performance of the covariate-only model was largely attributed to cohort differences; within cohorts, the covariate model was not predictive (UQ: Pearson’s R=8.48×10^-3^ p-value=0.95, UCSF: Pearson’s R=0.15, p-value=0.38) but epigenetics remained predictive (UQ: Pearson’s R=0.49, p-value=8.52×10^-4^ UCSF: Pearson’s R=0.54 p-value=7.24×10^-4^).

We also found that the epigenetic models predicting FVC and ALSFRS-R slope were also significantly better than covariate-only models (FVC p-value=2.67×10^-2^, ALSFRS-R slope p-value=4.10×10^-2^), but more mild than the ALSFRS-R models (FVC Pearson’s R=0.50, p-value=3.78×10^-3^, ALSFRS-R slope Pearson’s R=0.28, p-value=2.81×10^-2^) (Fig. 7b-c). Together, these results suggest that cfDNA epigenetic features are related to clinical traits used to measure ALS disease progression.

**Figure 7:**
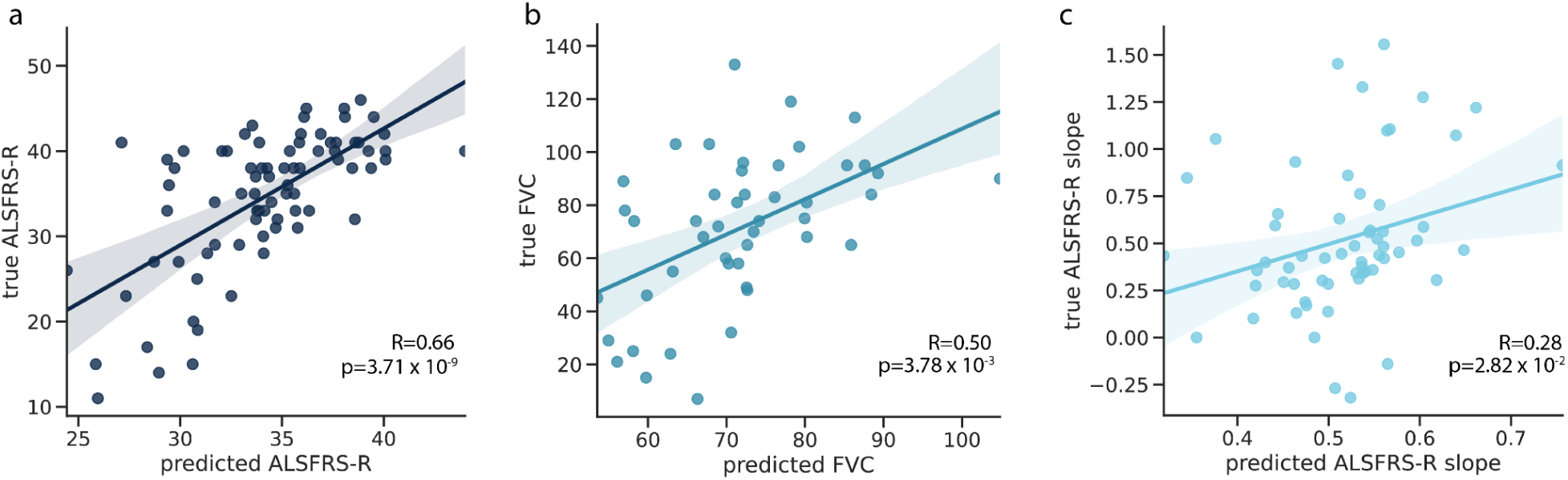
Predictive performance of cfDNA epigenetic features for ALS phenotypes. For a tenfold cross-validated model trained using cfDNA methylation proportion and coverage features the predicted versus true (a) ALSFRS-R, (b) FVC, and (c) ALSFRS-R slope. Each point represents one ALS case.

Lastly, we studied whether the same cfDNA epigenetic features that were associated with ALS disease phenotypes could differentiate between ALS and PLS cases. Due to the small sample number of PLS cases (n=15), we again combined the two cohorts and fit using 5-fold cross-validation with a non-penalized parameter for cohort. Although the analysis was underpowered, we observed a statistically significant difference between model predictions for ALS and PLS cases (AUC=0.74, linear regression effect size=36.61, p-value=1.9×10^-2^).

## 3 Discussion

Here, we presented a scalable cfDNA capture protocol that measures the methylation status of disease and tissue relevant CpG sites. We applied this capture technology to two independent cohorts of ALS patients and age-matched controls and examined the correlation with ALS disease status and progression. We then integrated both the read coverage and methylation proportion of the targeted sites in a machine-learning model. This model significantly discriminated between ALS patients and controls in two independent cohorts, including those with a variety of other neurological diseases. Together, our results suggest that a capture approach targeting tissue informative DNA methylation markers has value in quantitative biomarker development and that cfDNA has the potential to be a clinically relevant biomarker for ALS.

A key strength of using methylation markers informative of a broad variety of tissues is that it facilitates a comprehensive picture of a patient’s biological state and is not limited to a specific tissue or context. For example, neurofilament light chain is an exciting biomarker candidate for ALS.^49–51^ However, neurofilament light chain also is elevated in other neurodegenerative diseases, which might limit its specificity for some applications.^52^ By capturing and quantifying methylation levels at multiple tissue-informative CpG sites simultaneously, the panel has the potential to also learn about biological processes occurring in ALS outside of neurodegeneration. In particular, cfDNA is well-suited to measuring inflammation,^6,35^ which has been of recent interest in ALS pathophysiology.^37,39^ Future work could provide additional insight into how cfDNA relates to markers in ALS, providing a complementary avenue for investigation into disease mechanisms.

We observed differential performance between the UQ and UCSF cohorts. Specifically, the UCSF model outperformed the UQ model. Furthermore, the transferability was better when the UQ model was applied to the UCSF cohort. While it is likely a combination of factors, one explanation may be attributed to differences in sequencing depth. The UCSF cohort had higher on-target CpG coverage. Additional coverage may reduce noise, especially in analyses utilizing methylation proportion. In some cases, the overall coverage is limited by the total amount of cfDNA available as input to the sequencing assay. This could be improved by recent high-throughput extraction technologies with the ability to increase cfDNA yield from a plasma sample.^53,54^

Model performance also may be affected by the slight differences in ALS patient characteristics between the cohorts. For example, the UCSF cohort had patients with lower ALSFRS-R scores and whose advanced condition may be easier to detect in cfDNA. ALS is also an extremely heterogeneous disease,^55^ which can make designing biomarkers that generalize across patient populations difficult. It is also important to note that both cohorts were of majority European ancestry. Further exploration of how epigenetic cfDNA profiles differ between diverse subtypes of patients or change longitudinally as patients progress is now needed.

This study also only examined the performance of tissue informative markers in characterizing ALS. Since initiating these studies other proposed blood-based biomarkers for ALS, like neurofilaments,^52,56,57^ proteomics,^58^ or miRNA^13^ have demonstrated promise and future studies will need to benchmark with at least one of these. Previous studies have also illustrated the benefit of combining different types of biomarkers to enhance predictive performance. Future work on cfDNA biomarker development in ALS could assay multiple biofluids simultaneously and include a range of cohorts (i.e. asymptomatic gene-positive carriers for diagnosis, multi-ancestry, neurological conditions presenting with weakness). Integration of these multiple measurements, along with information about existing patient genetic liability, would robustly test its potential context of use and may improve disease prediction models.

Lastly, there are numerous avenues for improving algorithms associated with the approach outlined here. While methylation capture arrays allow for a more cost-effective and focused analysis over relevant CpG sites, targeted capture also limits the coverage of the genome. This has the potential to miss important methylation changes occurring outside the targeted regions. Additionally, since we relied on published tissue methylation data sets that are low coverage and inherently noisy, TIM selection might be affected. Marker selection and overall algorithm performance might be improved by better, high-coverage reference data. Reference panel design for cfDNA applications is a robust area of current research, and incorporating new samples or biobanks into ALS disease prediction could be an area for future research. Finally, single-molecule^59^ and nonlinear models^31^ have shown recent promise in the analysis of cfDNA profiles.

Overall, the design of the cell-free DNA methylation capture panel and related prediction algorithms presented in this study represents a significant advancement in the field of ALS research. They demonstrate promising potential as a non-invasive and diagnostic tool for ALS, which could facilitate timely intervention and personalized treatment strategies. Further research and validation are necessary to refine the panel’s performance, assess its generalizability, and address practical considerations. Nonetheless, this study paves the way for the integration of DNA methylation biomarkers into the clinical management of ALS, bringing us closer to improved patient outcomes.

## 4 Methods

### 4.1 Patient Recruitment and Clinical Data

A total of 192 participants were enrolled in a prospective manner at the UCSF ALS Clinic in San Francisco, California, USA, the Royal Brisbane and Women’s Hospital and Mater Hospital in Brisbane, Australia under neurologist supervision from 2018-2021. All participants provided written informed consent and the study received approval from the Human Research Ethics Committee at the Royal Brisbane and Women’s Hospital (HREC/17/QRBW/299) and by the UCSF Committee on Human Research (IRB 10-05027).

Patients (with ALS/being assessed for ALS) and when possible, control (non-related, closely age-matched family members, caregivers or volunteers) were recruited. A second set of other neurological controls were recruited from a non-ALS outpatient clinic under neurologist supervision. Allocation to diagnostic groups was performed according to the latest available clinical information (clinical censor date October 2024).

For cases and controls, age, sex, and self-identified race/ethnicity (SIRE) were recorded. For ALS cases at the time of visit, FVC and ALSFRS-R were taken, and ALSFRS-R slope and FVC slope relative to the previous visit were calculated. The symptom onset site and date of first symptoms were also recorded.

To stabilize the cell-free DNA, all blood samples were collected in the PAXgene Blood ccfDNA Tubes following a clinic appointment. To ensure enough cfDNA was available for downstream applications 20 mL of whole blood from controls/OND and 10 mL of whole blood from cases were collected. Following laboratory receipt (typically within 24-48hrs of collection) blood was spun with the brake off (10mins, 1900g) before plasma was aliquoted and spun twice (10mins, 16000g) to remove any further debris. Plasma was then stored at -80 before further processing.

### 4.2 Library Preparation and Sequencing

Using a harmonized protocol across two sites (UCSF and UQ) cfDNA was extracted and prepared for sequencing. Briefly, plasma was thawed at room temperature and cfDNA was extracted from all available plasma (range 2-8 ml) using the QIAGEN Circulating Nucleic Acid kit (Cat No: 55114) according to the manufacturer’s recommendations. Extracted cfDNA was quantified using Qubit dsDNA HS Assay and visualized using the cfDNA assay (Agilent - TapeStation 4200 (UCSF) and Agilent Bioanalyzer 2100 (HS kit) (UQ)). cfDNA was bisulfite converted using the Zymo Lightning kit (Zymo Research) and underwent library preparation using the Accel-NGS Methyl-Seq (Swift Biosciences) according to the manufacturer’s instructions with a major modification. Briefly, the denatured BS-converted cfDNA was subject to the adaptase, extension, and ligation reaction. Following the ligation purification, the DNA underwent primer extension (98C for 1 minute; 70C for 2 minutes; 65C for 5 minutes; 4C hold) using oligos containing random UMI and i5 barcodes. #The extension using a UMI-containing primer allows the tagging of each individual molecule in order to be able to remove PCR duplicates and correctly estimate DNA methylation levels.

Following exonuclease I treatment and subsequent purification, the libraries were then amplified using a universal custom P5 primer and custom i7-barcoded P7 primers (initial denaturation: 98C for 30 seconds; 15 cycles of: 98C for 10 seconds, 60C for 30 seconds, 68C for 60 seconds; final extension: 68C for 5 minutes; 4C hold). The resulting unique-dual indexed libraries were then purified, quantified using the Qubit HS-dsDNA assay, the quality checked using the D1000-HS assay (Agilent - TapeStation 4200), and grouped as 12-plex pools. Each pool was then subject to hybridization capture using the xGen Hybridization Capture Kit (IDT) using custom probes designed on approximately 5000 pre-selected regions.

For each top and bottom strands of the regions of interest, two probes were designed: one “unmethyl” probe with all G bases converted to A, and one “methyl” probe with all non-CpG G bases converted to A.

Following the hybridization capture, a final amplification PCR (initial denaturation: 98C for 30 seconds; 10 cycles of: 98C for 10 seconds, 60C for 30 seconds, 68C for 60 seconds; final extension: 68C for 5 minutes; 4C hold) has been performed, followed by SPRI beads purification and quantification as QC as previously described. To maximize consistency across sites, the same probes were used (shipped to Australia following UCSF library preparation).

The final pool of libraries was submitted for sequencing on an Illumina NovaSeq6000 (USA; UCSF Sequencing facility, Australia;UNSW Ramaciotti Sequencing facility) using identical run conditions (S4 lane - 150 PE, 8bases for i7, 17 bases for i5).

### 4.3 Tissue informative marker selection

TIMs were selected for 19 tissues and cell types: dendritic cells, endothelial cells, eosinophils, erythroblasts, macrophages, monocytes, neutrophils, T-cells, adipose, brain, fibroblast, heart, hepatocytes, lung, megakaryocytes, skeletal muscle, small intestine, placenta, and mammary epithelial cells. These tissues were determined based on our previous work to be relevant to ALS, or selected based on previous publications to be the primary contributors to cfDNA. At least two WGBS samples per reference dataset were obtained. The average methylation per CpG for the reference tissue replicates was calculated.

Per CpG, for one tissue at a time, the distance between the methylation proportion at that tissue and the mean methylation of all other tissues was calculated. The *N* sites per tissue with the greatest difference were kept as TIMs. If two tissues had the same CpG classified as a TIM, it was removed from both lists.

To begin, we selected 300 potential TIM sites and then performed quality control checks. To ensure that TIMs were sites that would be covered in cfDNA data, we used two WGBS cfDNA datasets and removed any CpG site that had less than an average of 10X coverage in both datasets. We also removed TIMs that overlapped a common SNP (minor allele frequency > 5%). Since we wanted to have the greatest diversity of regions targeted in the capture, if there were multiple TIM sites within 500bp of each other, we kept only the first site. Additional quality control was performed to remove TIMs overlapping repetitive regions and with low predicted target efficiency. After quality control, 4,994 TIMs remained.

### 4.4 Probe design

For each of the 4,994 TIMs, both a methylated and unmethylated probe were designed to bind to and capture both possible states of the targeted CpG. To increase the efficiency of the capture, 120 base pair probes were designed to target a window around the TIM. During bisulfite conversion, any cytosine base not protected by a methyl group in position 5 is converted into thymine.^60^ Since methylation in humans primarily occurs at CpG sites, this means that all cytosines on the forward strand would be converted to thymine. Thus, to capture the unmethylated CpG state, the unmethylated probe was designed with all guanine bases converted to adenine. For the methylated state, where only cytosines in a CpG dinucleotide would be protected from the bisulfite treatment, only non-CpG guanine bases were converted to an adenine.

### 4.5 Bioinformatic processing

For data generated at UCSF, UMIs were first extracted from the index read and added to the header of the corresponding R1 and R2 fastq file using umi_tools.^61^ This step was skipped for UQ samples since UMIs were not sequenced. For samples from both institutions, adapters were trimmed using trim_galore. Read alignment, processing, and methylation calling were performed using BsBolt v 1.6.1^62^ in an adapted pipeline published in Morselli et at.^18^ Reads were aligned to an hg38 bisulfite converted genome, which was generated using the BsBolt Index over an hg38 fasta file obtained from the UCSC genome browser. Reads were aligned using BsBolt Align in paired end mode with default parameters. To prepare for duplicate removal, aligned reads were subject to samtools fixmate and sorted.^63^ Umi_tools^61^ dedup in paired end mode was used to remove duplicate reads.

For both cohorts, CpG methylation was called using the command BsBolt CallMethylation-BG-CG-remove-ccgg. The CG parameter restricted to only CpG sites (ignoring non-CpG methylation), the the BG parameter sent the output to a bedgraph file and the -remove-ccgg parameter removed methylation calls in ccgg regions.

### 4.6 Genetic sex

As a quality control metric, we estimated the genetic sex of the samples and assessed how they corresponded to self-reported sex. We did this using scripts from Phung et al,^64^ which calculates the number of reads mapped to chromosome 19 and compares them to the number of reads mapped to the X chromosome. In individuals assigned female at birth, the ratio of chromosome 19 reads should be approximately 1 since they have two X chromosomes and two chromosome 19. We removed one individual whose genetic sex did not match their reported clinical data.

### 4.7 Deconvolution

cfDNA deconvolution was performed using CelFiE, which is a supervised deconvolution algorithm that is designed for noisy read count data and missing reference tissues. Input sites for CelFiE were the on-target TIMs selected for capture, As demonstrated in the CelFiE publication, summing reads from adjacent CpGs can improve deconvolution performance by decreasing sampling noise. As such, reads were summed +/-250bp around the target CpG. Sites with no reads covering the CpG were set to have a read depth of zero.

Deconvolution was performed using tissues representing organs and hematopoietic cell types, selected for their relevance in cfDNA.^10,28,29^ CelFiE can estimate an arbitrary number of unknown tissues. Since CelFiE learns from both the input and reference data, the number of samples influences the accuracy of unknown estimation. Based on simulation experiments published in the original CelFiE paper, 2 unknowns were chosen for the sample size of 96 total cfDNA input samples.

The reference panel for CelFiE consisted of 19 tissues over the same on-target TIMs as the input matrix. Reference samples were WGBS samples obtained from ENCODE^25^ and Blueprint.^26,65^ Reference samples were also summed in 500bp regions around the target CpG.

CelFiE was run over the UMI-deduplicated UQ samples, the UMI-deduplicated samples, and both cohorts combined. The CelFIE default of 10 random restarts was used.

After running deconvolution, differences in cell-type proportion between cases and controls were tested for one tissue at a time using the Python StatsModels package. A logistic regression model was run where the outcome was the binary case/control status and the input variable was the estimated tissue of origin proportion for a given tissue. Age, sex, and genetic ancestry were used as covariates.

### 4.8 Machine learning preprocessing

Samples with more than 10% of targeted CpGs missing, meaning that no reads were covering a CpG, were removed. Any site that had a median read coverage of 1 read or less was also removed. For the remaining sites and samples, the input matrix was made by dividing the number of methylated reads by the total number of reads. Imputation was performed per cohort over the methylation proportion matrix using SoftImpute, implemented in the Python package fancyImpute. For methylation coverage features, the coverage was normalized per sample by dividing the number of reads at a CpG by the total number of sequencing reads per individual.

Sex and SIRE were one-hot encoded and added as columns in the input matrix. Age, cfDNA starting concentration, and total cfDNA input were included as continuous covariates. Two separate matrices were kept, one for the ALS case/control status, and one for the methylation proportion and covariates.

### 4.9 Disease classification

Elastic net regression was performed in R using the BigStatsR package and big_spLogReg command.^42^ ALS disease status served as the binary outcome variable, while the DNA methylation proportion at targeted CpGs and clinical variables served as predictors. We incorporated age, genetic sex, SIRE, cfDNA concentration (nanograms/microliter), and total input cfDNA quantity (nanograms) into the regression models as non-penalized variables.

Models were first trained on each cohort separately and then applied to the second cohort. The alpha parameter which controls model sparsity, was selected by performing ten-fold cross-validation on the training cohort and picking the optimal value. The BigStatsR package removes the manual selection of an optimal lambda value by introducing the Cross-Model Selection and Averaging (CMSA) procedure.^42,66^ In brief, CMSA separates the training set into K folds and then performs cross-validation within the training set to obtain a set of vectors of predictions. This set of coefficients is averaged to produce the final coefficient value. For our model, we used the BigStatR default K value of 10. To standardize the weights produced per CpG site in each model, we scaled input value parameters to have mean zero and variance one. We scaled the test and training data separately.

Cohort-only models were trained only within a single cohort using ten-fold cross-validation. To evaluate the overall performance of the two cohorts, we trained a single model combining both sets of data and adding the cohort site as a non-penalized covariate. We used generalized linear models with a logit link function and additionally report area under the receiver operator curve (AUC).

### 4.10 Analysis of important features

To examine the importance of important DNA methylation and methylation coverage features in making model predictions, we obtained the weights, or β-values, at each feature from the combined cohort model. We merged the feature β-values with information on what tissue a TIM was selected for and whether it was hyper- or hypo-methylated. We used HOMER^67^ to intersect a TIM with the closest gene to the TIM site.

To assess the relationship between the methylation or coverage at a specific TIM site, we performed a logistic regression, with the methylation value of the samples as the predictor and case-control status as the outcome. We used SIRE, age, sex, cfDNA concentration, and total cfDNA input as covariates.

### 4.11 ALS disease phenotype prediction

ALS disease prediction models were trained for ALSFRS, ALSFRS Slope, and FVC. The top 1000 methylation features and top 1000 coverage features from the combined case-control prediction model were used as input to the model along with age, sex, SIRE, input cfDNA concentration, and total cfDNA input as non-penalized covariates. Due to low sample sizes for the case-only analysis, we meta-analyzed the two cohorts and additionally added cohort as a non-penalized covariate. We trained the elastic net model using the BigStatsR package with the big_spLinReg command. Each of the three models were evaluated against an elastic net model trained on only the covariates.

### 4.12 Off target prediction models

Off target prediction models incorporated information for all CpGs obtained from high throughput sequencing. To do this, we found the union of all sites across all samples in a cohort. To maximize the number of off-target sites considered, we then removed sites with more than 5% missingness. Due to the lower coverage and increased number of sites, we did not impute missing sites. Since cohorts had differences in sequencing depth and on-target coverage, sites were analyzed separately. Case control status was then predicted using ten fold cross validation in an elastic net model using the BigStatsR package in the same manner as the on-target models.

### 4.13 Downsampling simulations

To simulate samples with lower read depth, we used picard DownsampleSam^68^ to randomly remove reads at specified proportions of the total starting amount of reads to produce a bam file. We did this for each UCSF cfDNA sample. Then, methylation was re-called on the downsampled bam file using BsBolt to produce a new estimate of the methylation proportion and coverage of a CpG. We then subset to the on-target CpGs and individuals used in Fig.5b, imputing any missing values with SoftImpute. Then, an elastic net model was trained as in Section 4.9. The ten fold cross validated AUC was recorded for each set of downsampled samples.

## Data Availability

Fastq files and associated metadata for the samples are freely available at NCBI GEO. Tissue and cell-type WGBS data is freely available on the ENCODE Project and BLUEPRINT Epigenome Project data access portals.

## 6 Acknowledgements

We are extraordinarily grateful to the participants of this study and their families. We are also thankful for helpful conversations with M. Yamakawa.

We would like to thank the contributors and funders of SALSA-SGC (Sporadic ALS Australia-Systems Genomics Consortium) for supporting infrastructure to share clinical data used for the UQ samples. We gratefully acknowledge the funding support provided by the ALS Association, ALS Finding a Cure, the ALS Biomarker Collaboration, the UCSF Weill Award, the Australian National Health and Medical Research Council (NHMRC) and the Motor Neurone Disease Research Institute Australia (MNDRIA). C.C. was partially supported by T32NS048004.

## 7 Code Availability

Scripts to replicate analyses can be found at https://github.com/christacaggiano/cfdna-tims

## 9 Supplementary Figures

**Figure S1:**
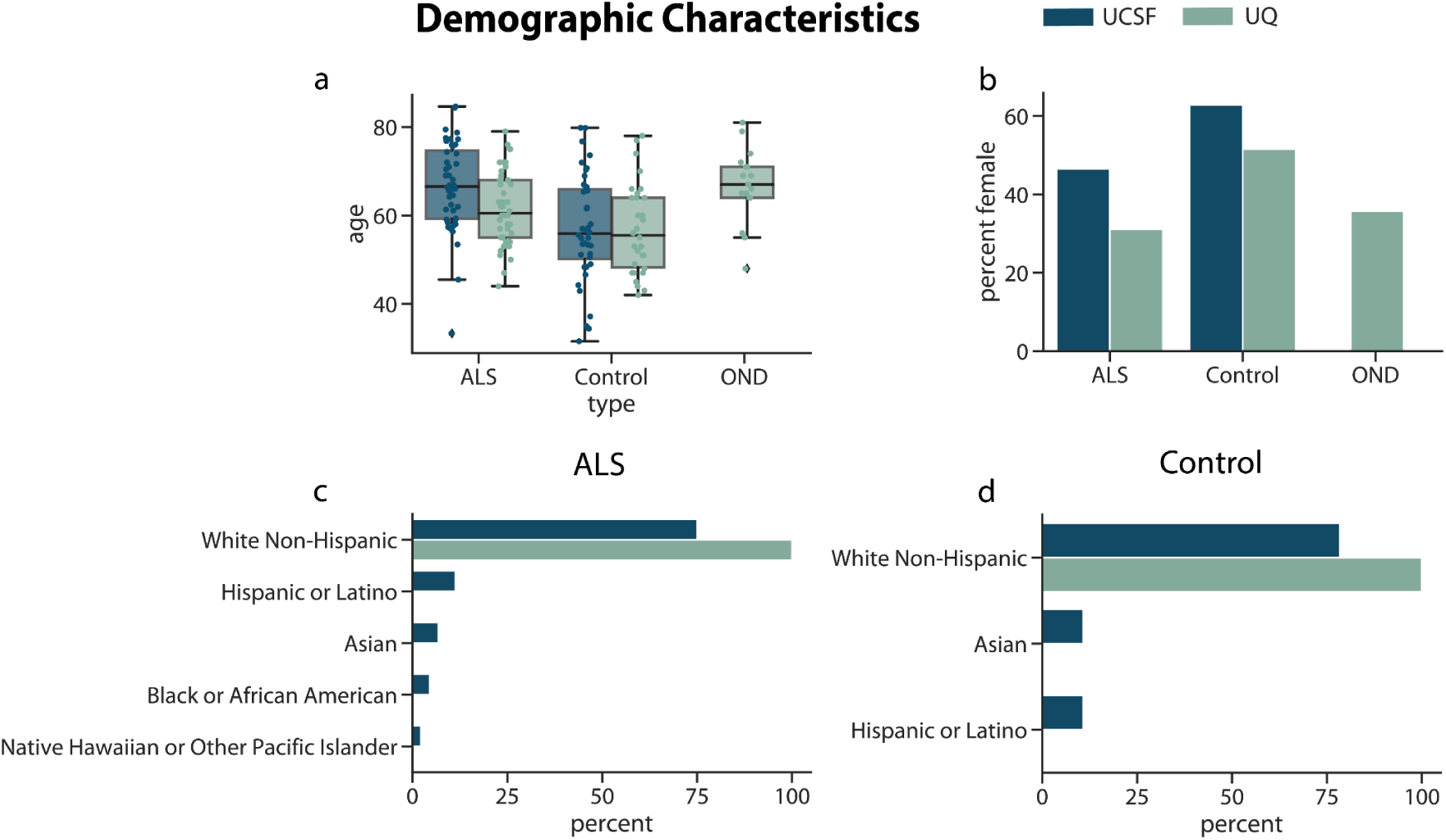
Cohort demographic characteristics. For the UQ and UCSF cohorts, (a) the distribution of the age of the cases and controls, (b) the percentage of the cohorts that are female, and the percentage of the (c) ALS cases and (d) controls that identify as five different racial/ethnic categories.

**Figure S2:**
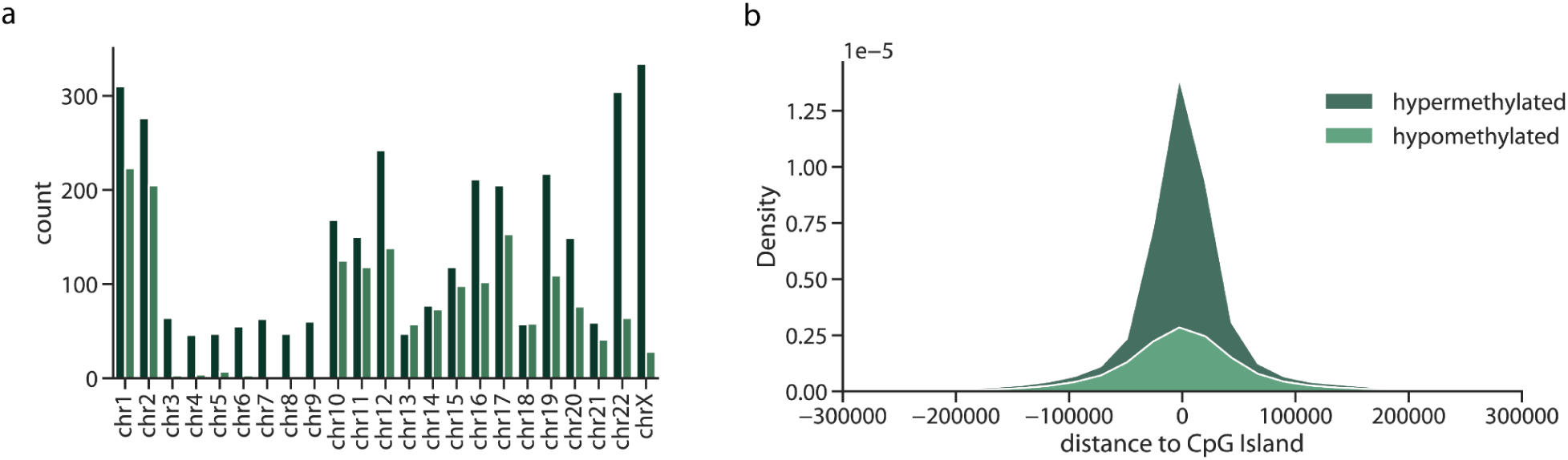
Properties of captured TIMs. **(a)** The number of TIMs selected per chromosome and (b) for the two types of TIMs, the distribution of distances between a TIM and a CpG island.

**Figure S3:**
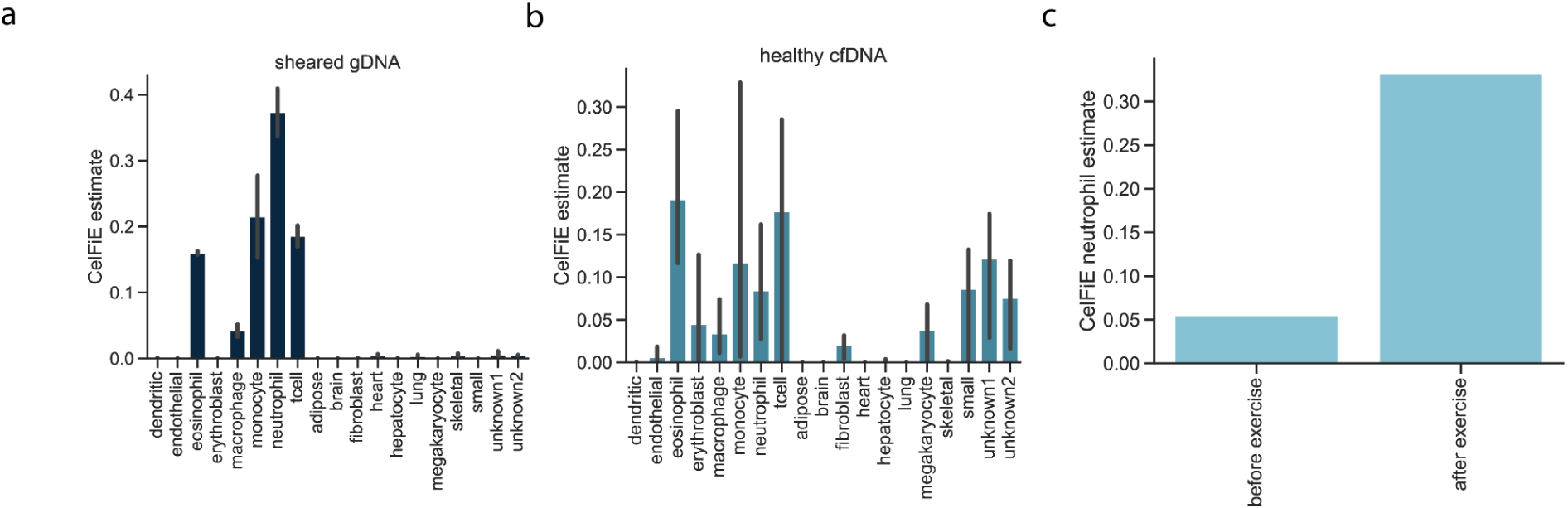
Deconvolution of validation data. The CelFiE estimates **(a)** for sheared genomic DNA (n=2) samples taken from blood and **(b)** healthy cfDNA (n=3). **(c)**For cfDNA taken from one individual before and after exercise, the proportion of cfDNA estimated to be originating from neutrophils.

**Figure S4:**
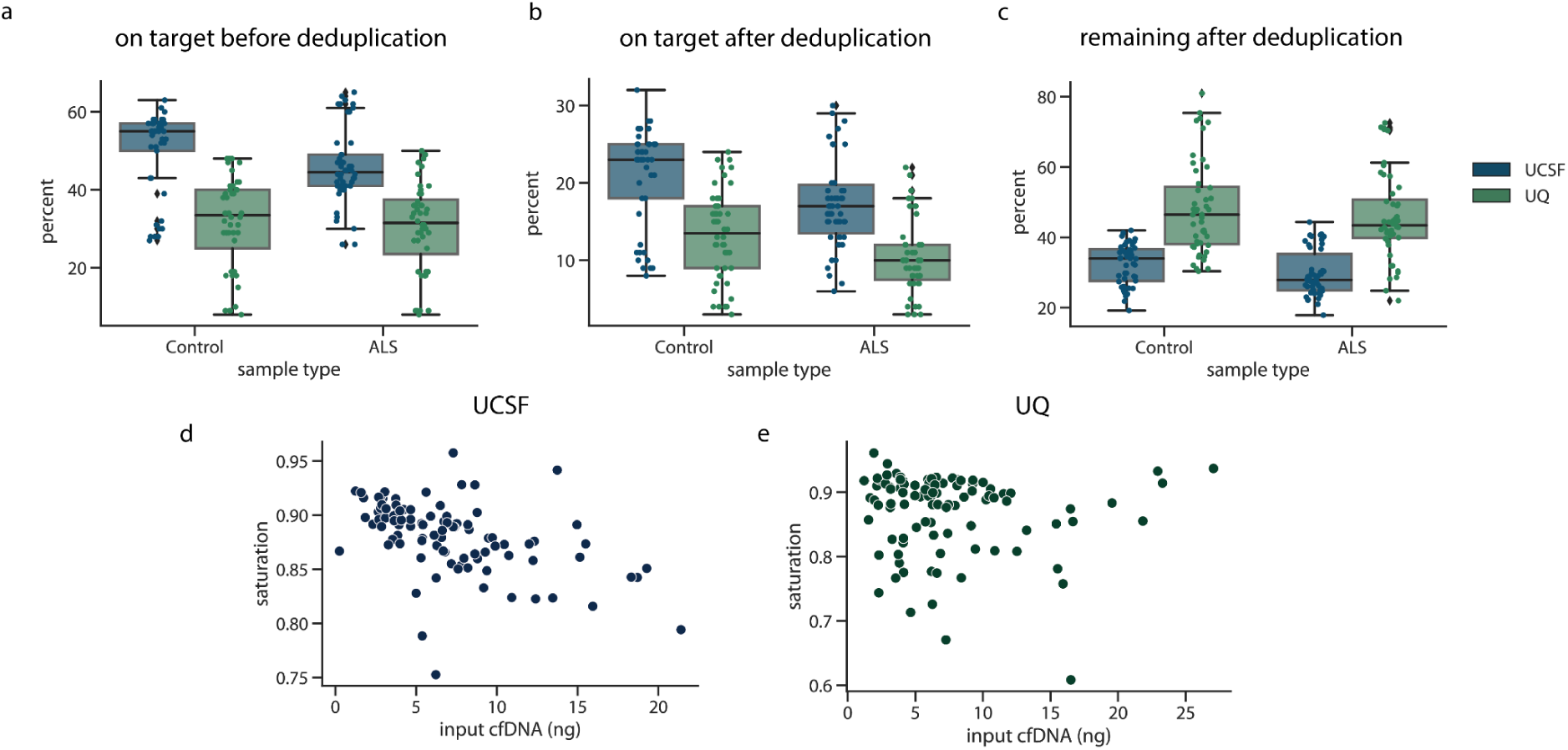
On target percentage. The percentage of reads that were on-target (**a)** before deduplication and (**b)** after deduplication. For each cohort, (**c)** the percentage of the total mapped starting reads before deduplication that remained after deduplication. The on-target saturation, defined as 1-(median depth on target after deduplication / median depth on target before deduplication) for (d) the UCSF cohort and (e) the UQ cohort.

**Figure S5:**
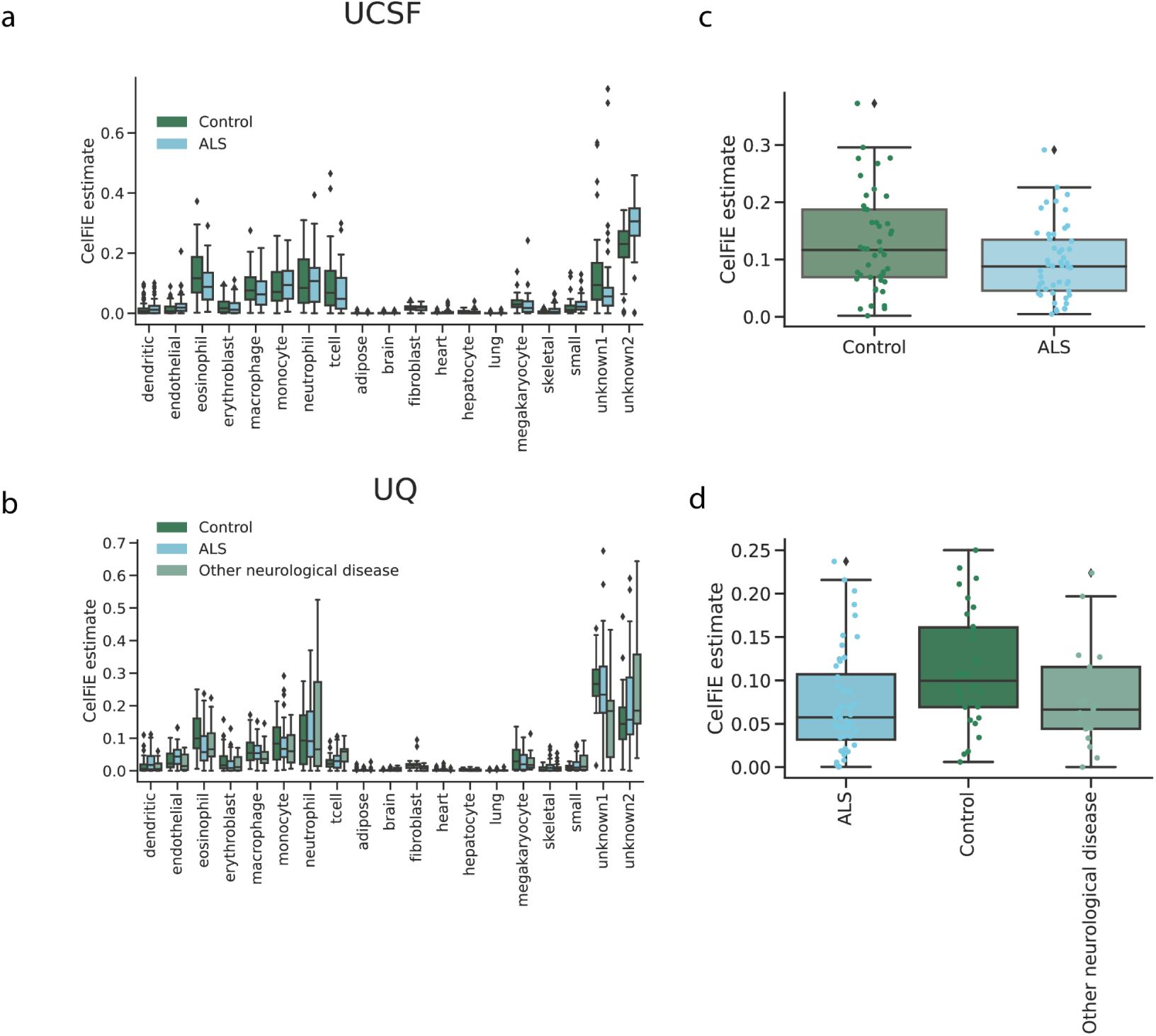
Cell-type decomposition estimates. The proportion of cfDNA estimated by CelFiE to originate from each tissue for each sample type in the (a) UCSF cohort and (b) UQ cohort.

**Figure S6:**
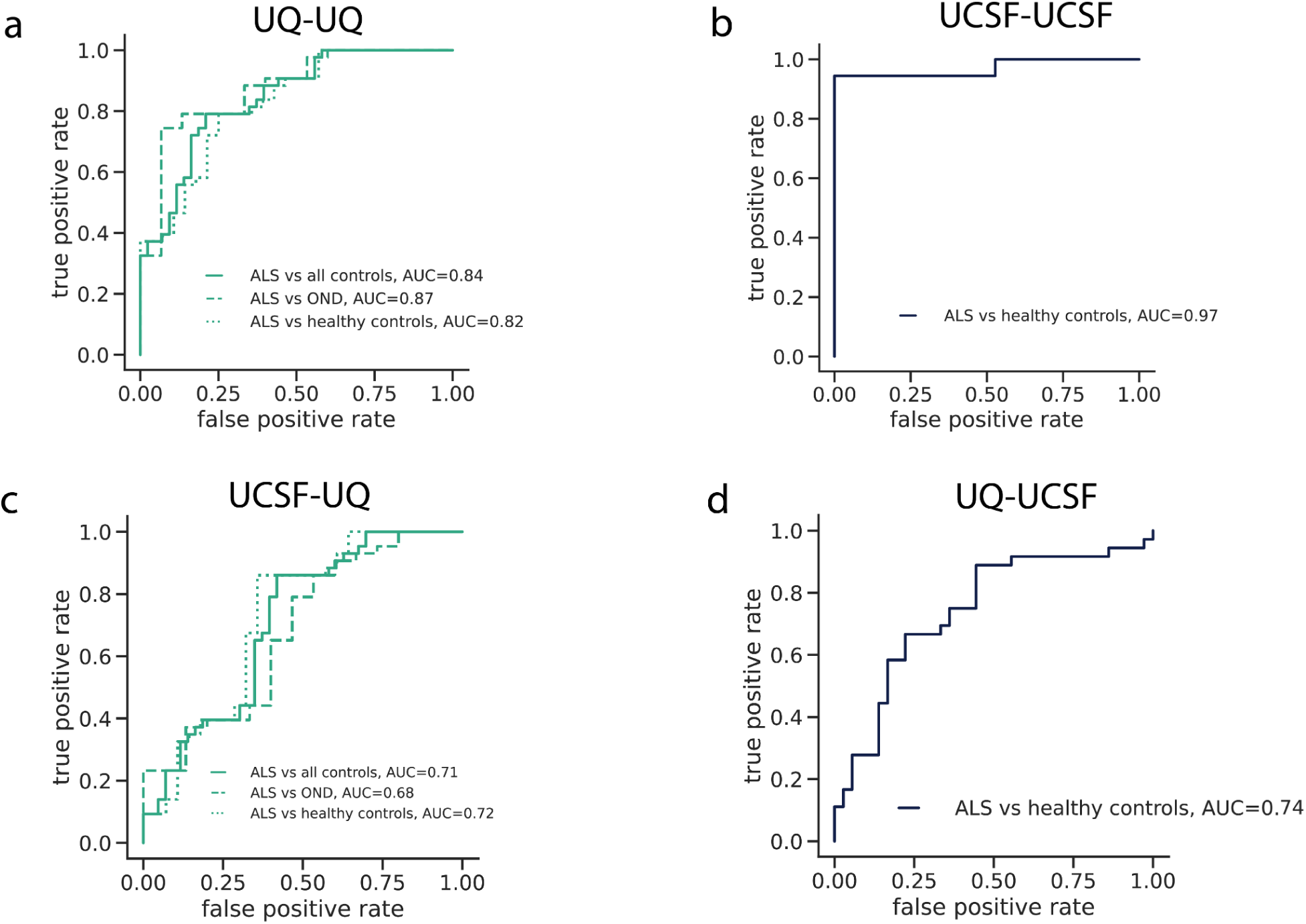
ALS classification using CpG coverage. The false positive rate versus true positive rate for models trained and tested using only CpG coverage as input features for (a) ten fold cross validation within UQ samples (b) ten fold cross validation within UCSF samples (c) trained on UCSF data and tested on UQ data, and (d) trained on UQ data and tested on UCSF data

**Figure S7:**
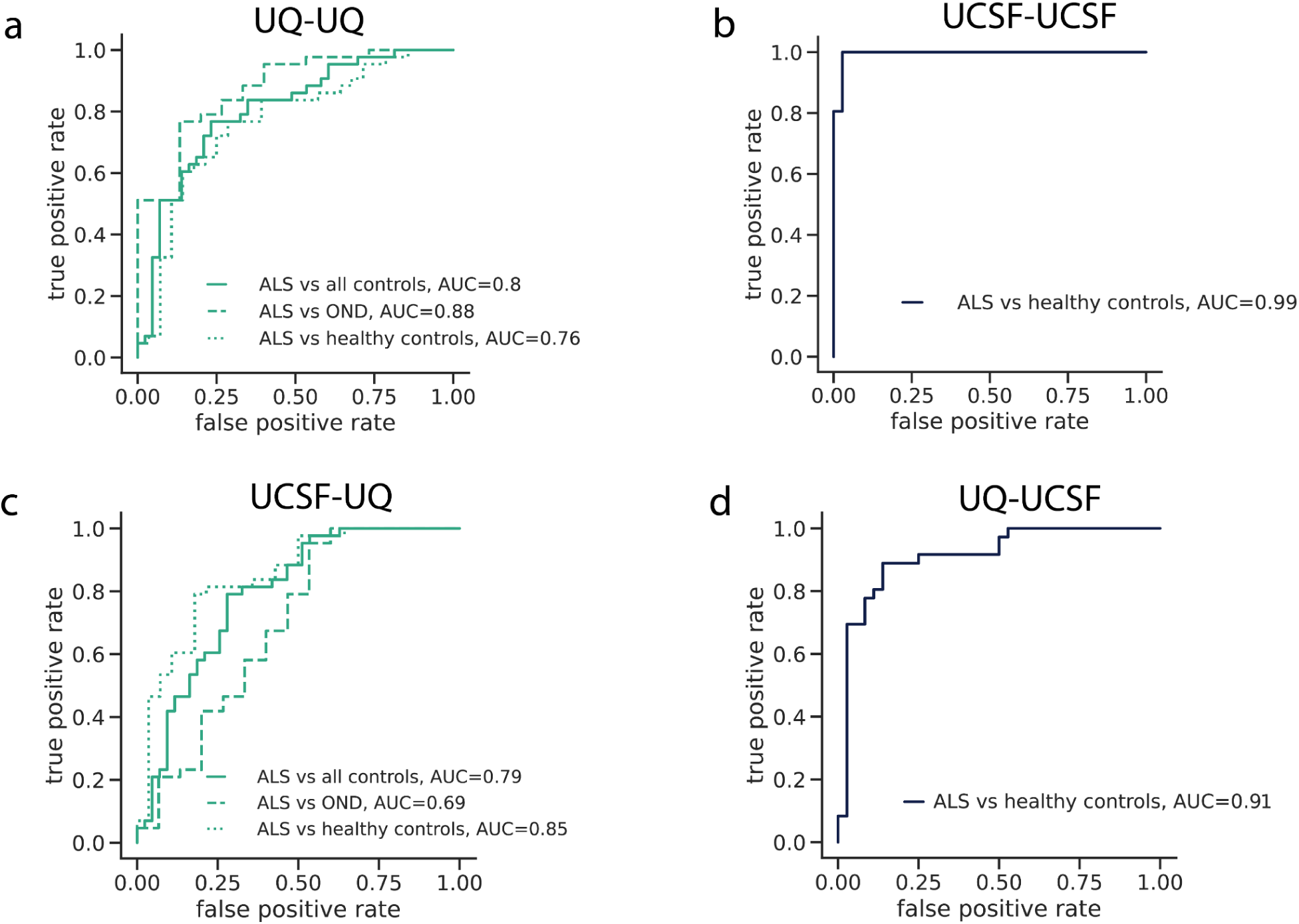
ALS disease classification using CpG methylation. The false positive rate versus true positive rate for models trained and tested using only CpG methylation proportion as input features for **(a)** ten fold cross validation within UQ samples **(b)** ten fold cross validation within UCSF samples **(c)** trained on UCSF data and tested on UQ data, and **(d)** trained on UQ data and tested on UCSF data

**Figure S8:**
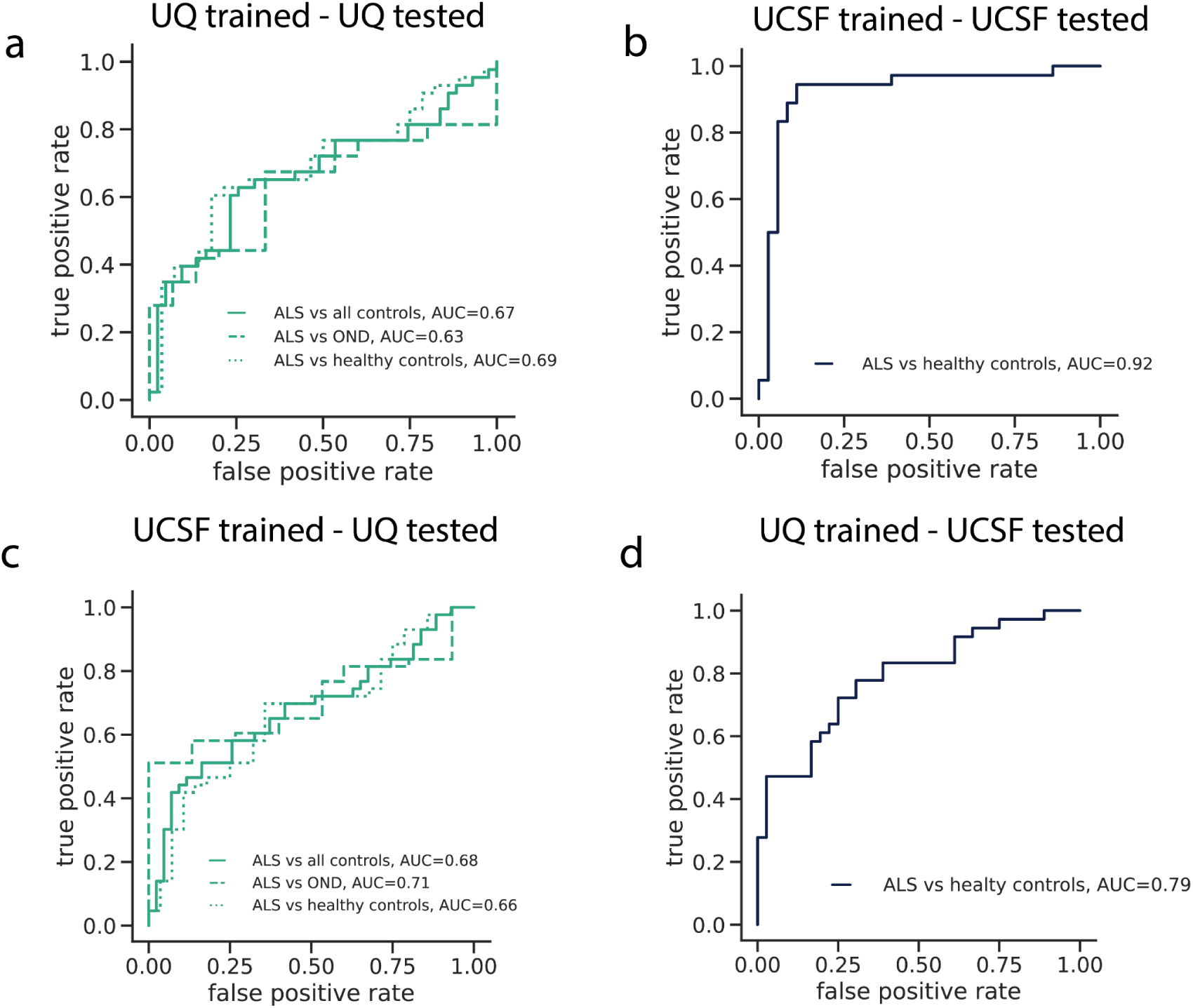
ALS disease classification using only covariate information. The false positive rate versus true positive rate for models trained and tested using only covariate information (age, sex, SIRE, starting cfDNA concentration, and total cfDNA input) as input features for **(a)** ten fold cross validation within UQ samples **(b)** ten fold cross validation within UCSF samples **(c)** trained on UCSF data and tested on UQ data, and **(d)** trained on UQ data and tested on UCSF data

**Figure S9:**
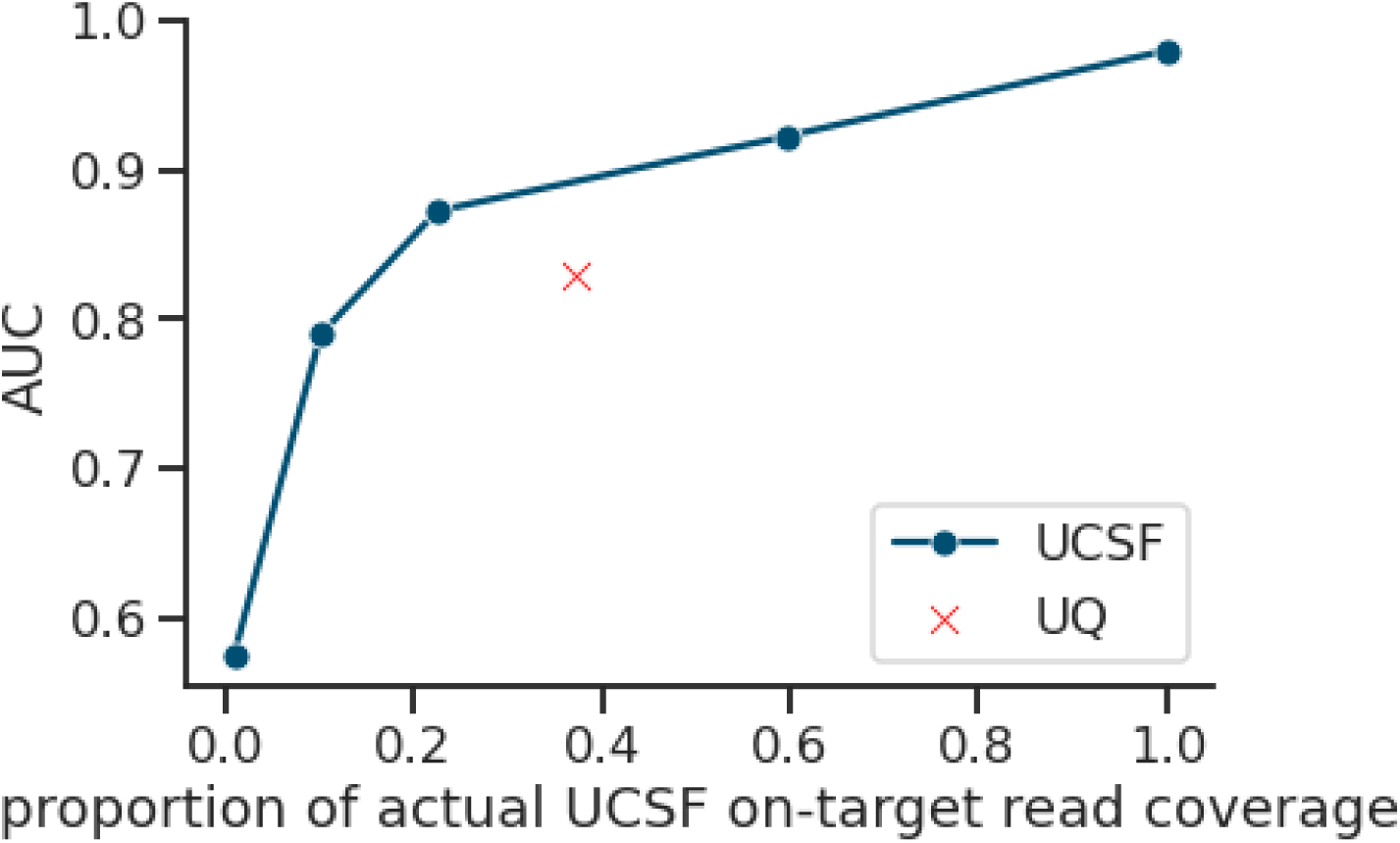
The relationship between read coverage and predictive performance. For UCSF cfDNA samples, the total number of reads was randomly downsampled to reduce overall on-target CpG coverage relative to the actual UCSF read coverage. The downsampled samples were then used as input for elastic net models trained using 10 fold cross validation to predict case-control status in the UCSF cohort and the AUC was recorded. The within-cohort UQ AUC is indicated by a red X.

**Figure S10:**
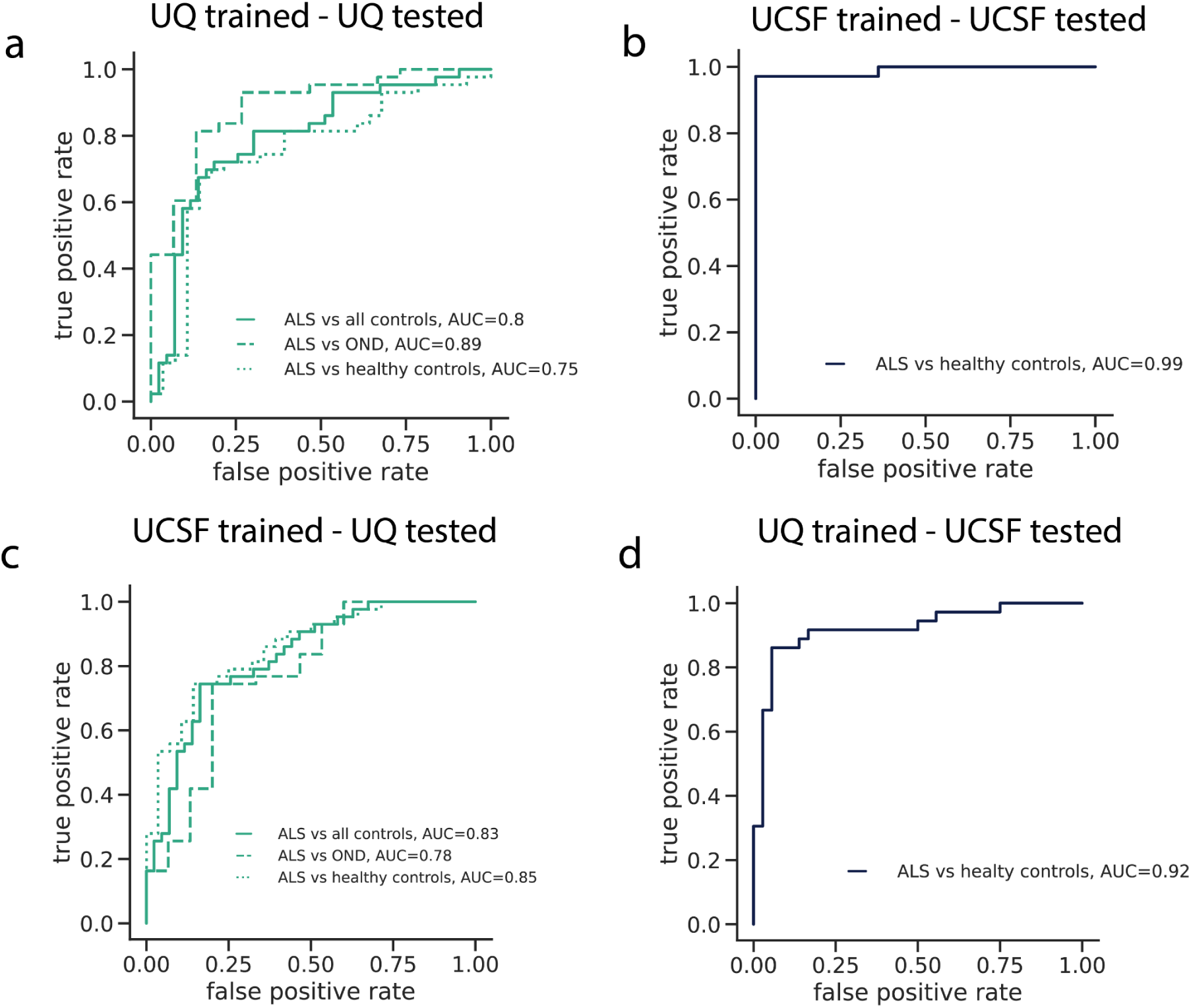
ALS disease classification without skeletal muscle TIMS. The false positive rate versus true positive rate for models trained and tested using cfDNA CpG methylation, CpG coverage, and covariate information (age, sex, SIRE, starting cfDNA concentration, and total cfDNA input) for all TIMs besides those chosen for skeletal muscle as input features for **(a)** ten fold cross validation within UQ samples **(b)** ten fold cross validation within UCSF samples **(c)** trained on UCSF data and tested on UQ data, and **(d)** trained on UQ data and tested on UCSF data

**Figure S11:**
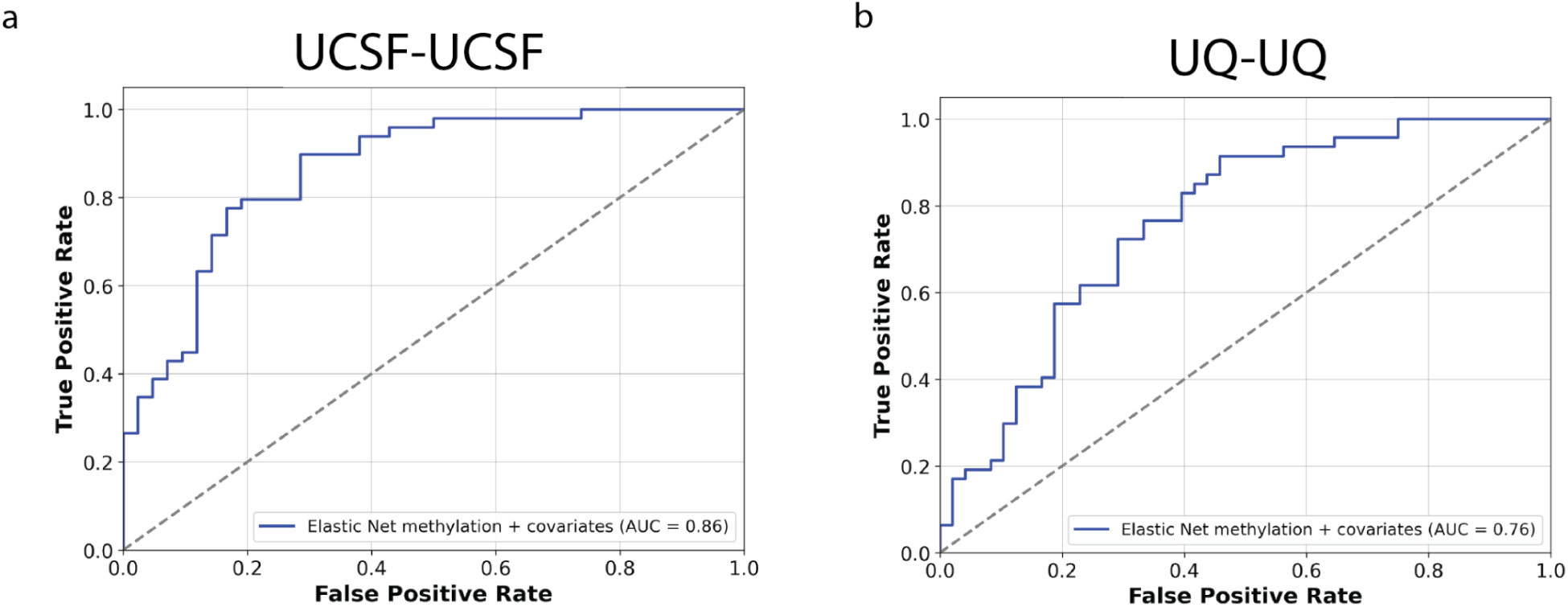
ALS disease classification with off-target CpGs. The false positive rate versus true positive rate for models trained and tested using off target cfDNA CpG methylation trained and tested used **(a)** ten fold cross validation within UCSF samples **(b)** ten fold cross validation within UQ samples.

## Notes

### Competing Interest Statement

The authors have declared no competing interest.

### Author Declarations

All participants provided written informed consent and the study received approval from the Human Research Ethics Committee at the Royal Brisbane and Women's Hospital (HREC/17/QRBW/299) and by the University of California San Francisco Committee on Human Research (IRB 10-05027).

